# Correlation of patient serum IgG, IgA and IgM antigen binding with COVID-19 disease severity using multiplexed SARS-CoV-2 antigen microarray and maintained relative IgA and IgM antigen binding over time

**DOI:** 10.1101/2022.08.22.22278930

**Authors:** Marie Le Berre, Terézia Paulovčáková, Carolina De Marco Verissimo, Seán Doyle, John P. Dalton, Claire Masterson, Eduardo Ribes Martínez, Laura Walsh, Conor Gormley, John G. Laffey, Bairbre McNicholas, Andrew J. Simpkin, Michelle Kilcoyne

**Affiliations:** Carbohydrate Signalling Group, Discipline of Microbiology, School of Chemical and Biological Sciences, University of Galway, University Road, Galway, H91 TK33, Ireland; Molecular Parasitology Lab, Centre for One Health and Ryan Institute, School of Natural Science, University of Galway, University Road, Galway, H91 TK33, Ireland; Department of Biology, Maynooth University, Maynooth, Co. Kildare, Ireland; School of Medicine, and Regenerative Medicine Institute (REMEDI) at CÚRAM Centre for Research in Medical Devices, University of Galway, Galway, Ireland; Lambe Institute for Translational Research, School of Medicine, College of Medicine, Nursing and Health Sciences, University of Galway, Galway, Ireland; Gene Center, Ludwig-Maximilians University of Munich, Feodor-Lynen-Str. 25, 81377 Munich, Germany; University College Dublin, Belfield, Dublin 4, D04 V1W8, Ireland; Royal College of Surgeons in Ireland, 123 St. Stephen’s Green, Dublin 2, Ireland; Department of Anaesthesia and Intensive Care Medicine, University Hospital Galway, Saolta University Hospital Group, Newcastle Road, Galway, Ireland; School of Mathematical and Statistical Sciences, University of Galway, University Road, Galway, H91 TK33, Ireland

## Abstract

Zoonotic spillover of severe acute respiratory syndrome coronavirus 2 (SARS-CoV-2) to humans in December 2019 caused the coronavirus disease 2019 (COVID-19) pandemic. Serological monitoring is critical for detailed understanding of individual immune responses to infection and protection to guide clinical therapeutic and vaccine strategies. We developed a high throughput multiplexed SARS-CoV-2 antigen microarray incorporating spike (S) and nucleocapsid protein (NP) and fragments expressed in various hosts which allowed simultaneous assessment of serum IgG, IgA, and IgM responses. Antigen glycosylation influenced antibody binding, with S glycosylation generally increasing and NP glycosylation decreasing binding. Purified antibody isotypes demonstrated a binding pattern and intensity that differed from the same isotype in the presence of other isotypes in whole serum, probably due to competition. Using purified antibody isotypes from naïve Irish COVID-19 patients, we correlated antibody isotype binding to different panels of antigens with disease severity, with significance for binding to the S region S1 expressed in insect cells (S1 Sf21) for all three antibody isotypes. Assessing longitudinal response for constant concentrations of antibody isotypes for a subset of patients demonstrated that while the relative proportion of antigen-specific IgGs decreased over time for severe disease, the relative proportion of antigen-specific IgA binding remained at the same magnitude at 5 and 9 months post-first symptom onset. Further, the relative proportion of IgM binding decreased for S antigens but remained the same for NP antigens. This may support antigen specific serum IgA and IgM playing a role in maintaining longer-term protection, of importance for developing and assessing vaccine strategies. Overall, these data demonstrate the multiplexed platform as a sensitive and useful platform for expanded humoral immunity studies, allowing detailed elucidation of antibody isotypes response against multiple antigens. This approach will be useful for monoclonal antibody therapeutic studies and screening of donor polyclonal antibodies for patient infusions.

## 1. Introduction

Severe acute respiratory syndrome coronavirus 2 (SARS-CoV-2) is the viral causative agent of the current global pandemic of coronavirus disease 2019 (COVID-19). Disease symptoms vary in severity in individuals, ranging from no symptoms to pneumonia, and can result in SARS, acute respiratory distress syndrome (ARDS), and death. As of 5^th^ August 2022, there have been over 579 million confirmed COVID-19 cases and over 6.4 million deaths worldwide due to the disease [1]. The majority of SARS-CoV-2-infected individuals produce specific serum antibodies by 1-3 weeks post-first symptom onset (PFSO), with immunoglobulin M (IgM) appearing typically in the first week, followed by IgG by the second week, and IgA appearing between the first and second week, although there is a large variety in individual timelines reported [2-6]. Serum antibodies generally reach maximum concentration by week 3-5 PFSO, but reports vary depending on individual response, serological testing method, and disease severity [2, 7]. To achieve protective immunity, antibodies produced against the pathogen must target specific viral proteins.

The SARS-CoV-2 enveloped virus expresses various proteins including membrane (MP), envelope, and spike (S) proteins on the envelope surface and the nucleocapsid protein (NP) located inside the virus particle [8]. The virus genome also contains two proteases, a papain-like protease and a 3C-like protease (3CLike). Serum antibody isotypes can develop against any viral antigen and an individual usually develops varying immune responses against a collection of presented antigens including S and NP proteins. The various viral antigens are glycosylated *in vivo* and their glycosylation contributes to immune recognition and binding interactions [9], though antigens produced in various recombinant systems and used in assays and diagnostics may have different or no glycosylation. The S protein consists of two regions, S1 and S2, and the S1 subunit contains the receptor binding domain (RBD), which is an important target antigen for an effective immune response. When the S glycoprotein is assembled as a trimer on the virion surface, the RBD binds to the angiotensin-converting enzyme 2 (ACE2) receptors on human cell surfaces to initiate infection [8]. Antibodies which bind to the viral RBD or S1 proteins can block the RBD-ACE2 interaction in the respiratory tract, stopping infection, and are known as the most potent neutralising antibodies [10, 11]. Neutralising RBD-specific IgG antibodies are associated with protection from re-infection for up to 6 months and decreased disease severity upon breakthrough infection [12, 13]. Higher and earlier titres of antibody isotypes binding to specific antigens and neutralising antibodies have been reported for patients with severe COVID-19 disease along with longer duration of antibody detection in the serum compared to those with mild disease [14-17]. It is of critical importance to monitor and understand the humoral immune response in relation to disease severity, protective immunity, duration of protection, and as a potential contributor to antibody-mediated immunopathology.

Real-time reverse transcription quantitative polymerase chain reaction (RT-qPCR) of viral material from nasopharyngeal swabs is considered the gold standard of diagnosis of SARS-CoV-2 infection. However, the reliability of this test is reduced in the first few days after exposure and in later days PFSO, as well as in younger patients and women for whom viral loads are usually lower [18-20]. Serological assays measuring serum antibodies against viral antigens can be used diagnostically to expand the detection window [21]. These assays are more typically employed to monitor immune response in individuals for disease surveillance, epidemiological studies, and monitoring vaccine response and efficacy, as well as assessing suitability of convalescent plasma donors for parenteral antibody treatments, and monoclonal antibody clinical therapies. SARS-CoV-2 NP, S and RBD are the most widely used target antigens for COVID-19 serological assays [7, 22, 23]. Early antigen-specific antibodies were reported for the majority of COVID-19 patients [24] and accurate serological monitoring could also contribute to personalised strategies of patient management and optimising vaccination approaches. The enzyme-linked immunosorbent assay (ELISA) presenting one antigen to measure antibody binding is the most used clinical serological assay format but can suffer from sensitivity limitations depending on the selected sample preparation, detection, and quantification methods [25]. To facilitate high throughput and sensitive serological monitoring, a miniaturised and multiplexed accurate platform is needed, ideally one which can assess IgG, IgM and IgA isotypes against multiple antigens at the same time.

Several multiplexed SARS-CoV-2 antigen assay platforms have been developed recently, ranging from bead-based assays to multiplexed antigens conjugated to functional glass or gold surfaces and using fluorescence- or plasmon-based detection [26-34]. Most of these formats were employed to detect antigen binding of up to two antibody isotypes, IgG and IgM or IgG and IgA, and all used a dilution of serum or plasma as the serum antibody source with detection of isotype binding using a fluorescently labelled anti-isotype secondary antibody. This approach relies on the selectivity of the anti-isotype secondary antibody, but cross-reactivity of these detection antibodies between antibody isotypes is possible. Binding quantification and the relative pattern of interactions may also be influenced by competition between antibody isotypes for the same antigen [26]. Further, the use of a serum dilution as the source of antibodies in an assay discounts the confounding impact of the absolute concentration of antibodies in the serum. The latter is of particular concern for studies evaluating kinetic or longitudinal immune response considering that the absolute concentrations of antibodies in serum declines over time post-infection.

In this study, we developed a multiplexed SARS-CoV-2 protein antigen microarray incorporating NP and full length S proteins, as well as S protein domains (S1, S2, and RBD) expressed separately and in various systems, which resulted in antigens with different glycosylation. We purified IgG, IgM and IgA from serum obtained from mild, medium, and severe COVID-19 patients and from healthy pre-pandemic donors to obviate the reliance of anti-isotype detection antibodies. We used the multiplexed SARS-CoV-2 antigen microarray to simultaneously profile and quantify the binding specificity of constant concentrations of IgG, IgM, and IgA. In using these approaches, we excluded the impact of declining serum antibody titre over time and the reliance on anti-isotype antibody selectivity. We correlated the severity of COVID-19 disease response with specific serum antibody isotype binding to certain antigens, examined the impact of antigen glycosylation on antibody binding, and demonstrated changes in antibody recognition and binding over time. In addition, we also compared the immune response of vaccinated naïve donors with infected patients.

## 2. Materials and methods

### 2.1. Materials

Invitrogen™ IgG (total), IgM (total), and IgA (total) human uncoated ELISA kits (cat. nos. 88-50550-88, 88-50620-88, and 88-50600-88 respectively), NuPAGE 4-12% Bis-Tris gels, MOPS running buffer, Pierce™ bicinchoninic acid (BCA) protein assay kit, Nab™ Protein G 0.2 mL spin columns, POROS CaptureSelect™ IgM affinity matrix, Pierce™ Spin Columns, Snap Cap, Alexa Fluor™ 555-(AF555-)labelled goat anti-human IgG (2 mg/mL; cat. no. A21433), TRITC-labelled goat anti-human IgM (1 mg/mL; cat. no. A18840), TRITC-labelled goat anti-human IgA (1 mg/mL; cat. no. A18786), isopropyl-β-D-1-thiogalactopyranoside (IPTG), and AF555 NHS ester (succinimydyl ester) were purchased from Thermo Fisher Scientific Inc. (Dublin, Ireland). Peptide M agarose matrix for IgA purification was obtained from InvivoGen Europe (Toulouse, France). Amicon® Ultra 0.5 mL centrifugal filters in 3, 30, and 100 kDa molecular weight cut off (MWCO) were from Merck-Millipore (Cork, Ireland). A selection of SARS-CoV-2 protein antigens recombinantly expressed in human embryonic kidney 293 (HEK293) cells, insect (Sf21) cells, or *Escherichia coli* (Table 1 and S1) were purchased from RayBiotech (RB; Peachtree Corners, GA, U.S.A.) or R&D Systems (R&D; Bio-Techne, Abindon, U.K.). Nexterion® slide H microarray slides were supplied by Schott AG (Mainz, Germany). TRITC-labelled lectins were purchased from EY Laboratories, Inc. (San Mateo, CA, USA). Mouse monoclonal anti-6X His IgG labelled with CF™ 640R antibody (1 mg/mL), Luria Bertani (LB) broth, and Sarkosyl was from Sigma-Aldrich Co. (Dublin, Ireland). All other reagents unless indicated were from Sigma-Aldrich Co. and were of the highest grade available.

**Table 1.** Recombinant SARS-CoV-2 proteins used to construct the antigen (Ag) microarray, the Ag code, molecular mass (Mr) observed by SDS-PAGE, expression system, protein sequence expressed, supplier, catalogue number, and approximate print concentration (mg/mL). MPL, Molecular Parasitology Lab; RB, RayBiotech; R&D, R&D Systems. * Ag failed to conjugate and omitted from data.

| Ag code | Protein Ag | Mr (kDa) | Expression | Sequence | Supplier | Cat. no. | Print conc (mg/mL) |
| --- | --- | --- | --- | --- | --- | --- | --- |
| Npro Full Ecoli | Nucleocapsid protein, full length | 50 | *E. coli* | 2-1269 | MPL | - | 0.06 |
| 3CLike Ecoli | 3C-like protease | 34 | *E. coli* | 1-306 | MPL | - | 0.24 |
| Spro Ecoli | Spike protein, full length | 135 | *E. coli* | 10-1282 | MPL | - | 0.12 |
| S1Frag Ecoli | Spike protein fragment 1 | 75 | *E. coli* | 10-686 | MPL | - | 0.15 |
| S2Frag Ecoli | Spike protein fragment 2 | 54 | *E. coli* | 687-1283 | MPL | - | 0.06 |
| S2Pri Ecoli | Spike protein fragment 2 prime region | 38 | *E. coli* | 816-1283 | MPL | - | 0.01 |
| NP Ecoli | Nucleocapsid protein | 50 | *E. coli* | Met1-Ala419 | RB | 230-01104 | 0.95 |
| NP HEK | Nucleocapsid protein | 50-60 | HEK293 | Met1-Ala419 | RB | 230-30164 | 0.6 |
| NP Sf21 | Nucleocapsid protein | 44-53 | Sf21 | Met1-Ala419 | R&D | 10474-CV | 0.5 |
| S1 HEK | S1 subunit protein | 106-121 | HEK293 | Val16-Pro681 | R&D | 10569-CV | 0.5 |
| S1 Sf21 | S1 subunit protein | 78-92 | Sf21 | Val16-Pro681 | R&D | 10522-CV | 0.2 |
| S1 Full HEK | S1 subunit protein, full length | 120 | HEK293 | Val16-Gln690 | RB | 230-30161 | 0.45 |
| B117 RBD HEK | B.1.1.7 receptor binding domain | 34-38 | HEK293 | Arg319-Phe541 | R&D | 10730-CV | 0.25 |
| MP Ecoli* | Membrane glycoprotein, C-terminal | 15 | *E. coli* | Arg101-Gln222 | RB | 230-01124 | 0.17 |

### 2.2. SARS-CoV-2 protein antigen expression and purification in *E. coli*

Full length SARS-CoV-2 nucleocapsid protein (Npro Full Ecoli), spike protein (Spro Ecoli), spike protein fragment 1 (S1Frag Ecoli), spike protein fragment 2 (S2Frag Ecoli), and the spike protein fragment 2 prime region (S2Pri Ecoli) (Table 1 and S1) were recombinantly expressed in *Escherichia coli* BL21 and purified as previously described [21]. Briefly, the protein sequences were codon optimized for expression and cloned into the pET-28a(+) vector, and into pET-19b for nucleocapsid protein (Npro Full Ecoli; Genscript Biotech). The synthesized vectors were transformed into *E. coli* BL21 competent cells (ThermoFisher Scientific) and clones grown in LB broth supplemented with 50 μg/mL kanamycin, or 100 μg/mL ampicillin for Npro Full Ecoli, at 37 °C to OD_600nm_ of 0.6. Protein expression was induced with 1 mM IPTG for 4 h at 30 °C or with 0.5 mM IPTG for 3 h at 37 °C for Npro Full Ecoli.

For recombinant protein purification, bacterial pellets were treated with 0.1 mg/mL lysozyme in the presence of 40 mM dithiothreitol (DTT) for 1 h on ice. Proteins in inclusion bodies were solubilised as described by Schlager, *et al*. [35], using 1% (*w/v*) sodium dodecyl sulfate (SDS) buffer (8 mM Na_2_HPO_4_, 286 mM NaCl, 1.4 mM KH_2_PO_4,_ 2.6 mM KCl, 1% (w/v) SDS, pH 7.4) containing 0.1 mM DTT. After sonication (twice for 2 min each, 40% amplitude), samples were centrifuged (15,000 x *g*, 4 °C, 30 min), filtered (0.45 μm syringe filters), and the supernatant containing soluble recombinant protein was passed through a pre-equilibrated Ni-NTA beads column (Qiagen). The column was washed with 30 mL wash buffer (8 mM Na_2_HPO_4_, 286 mM NaCl, 1.4 mM KH_2_PO_4,_ 2.6 mM KCl, 0.1% Sarkosyl (w/v), 40 mM imidazole, pH 7.4), and recombinant protein eluted with 4 mL elution buffer (8 mM Na_2_HPO_4_, 286 mM NaCl, 1.4 mM KH_2_PO_4,_ 2.6 mM KCl, 0.1% Sarkosyl (w/v), 250 mM imidazole, pH 7.4). The purified protein was buffer-exchanged into phosphate buffered saline, pH 7.4, (PBS) containing 0.05% Sarkosyl. The soluble recombinant 3C-like protease within the supernatant was purified and dialysed using the Profinia Affinity Chromatography Protein Purification System (Bio-Rad), with the Bio-Scale™ mini Profinity™ IMAC and mini Bio-Gel P-6 desalting cartridges (Bio-Rad).

Recombinant soluble Npro Full Ecoli was extracted from *E. coli* by sonicating twice (2 min each, 20% amplitude) in lysis buffer (50 mM Tris, 100 mM NaCL, 1 mM EDTA, 10% (v/v) glycerol pH 8.0, with 1 mM phenylmethylsulfonyl fluoride (PMSF) and 4 μg/mL leupeptin), followed by centrifugation and dialysis into 20 mM H_2_NaPO_4_, 500 mM NaCl, 20 mM imidazole, pH 7.4. Protein was centrifuged, filtered (0.45 μm), applied to HisTrap HP columns (GE Healthcare) equilibrated in the same buffer, and eluted with 20 mM H_2_NaPO_4_, 500 mM NaCl, 500 mM imidazole, pH 7.4. Npro Full Ecoli was stored in the elution buffer.

Protein concentrations were estimated by Bradford protein assay (Bio-Rad) and the proteins visualised on 4-20% sodium dodecyl sulfate-polyacrylamide gel electrophoresis (SDS-PAGE) gels (Bio-Rad) stained with Biosafe Coomassie (Bio-Rad) to verify purity.

### 2.3. Serum samples

Sera were obtained from 26 patients of Irish ethnicity admitted to hospital presenting with COVID-19 symptoms 5-34 days post-symptom onset (PFSO) who tested SARS-CoV-2 positive by qRT-PCR (Table 2). These patients had not been previously exposed to SARS-CoV-2 and samples were collected during the first disease surge in Ireland. Serum was collected from two healthy donors who had never had any COVID-19 symptoms and were presumed to have not been previously infected with SARS-CoV-2 30 days after their second dose of the Pfizer-BioNTech COVID-19 mRNA (COMIRNATY) vaccine (administered 30 days after their first dose with the same vaccine). Ethical approval for the use of human serum samples was granted by the Galway University Hospital research ethics committee (C.A. 1928) and the National Research Ethics Committee (research permit 20-NREC-COV-20). All participants provided written informed consent prior to the study or assent followed by informed consent once able for patients admitted to the Intensive Care Unit (ICU). Samples were stripped of any identifying information, aliquoted, and assigned anonymised identification numbers by the biobank co-ordinator (BMcN). The patient information associated with the identification numbers are only known to the biobank co-ordinator. Negative control serum samples collected in 2018 before the COVID-19 pandemic were gifted by the Irish Blood Transfusion Service (Table 2). These samples were stored at -20 ºC before thawing and immunoglobulin fractionation.

**Table 2.** Serum sample information including biobank code, COVID-19 disease status of individual at sampling time (mild, moderate, severe, or non-COVID-19 (NC)), days post-symptoms onset at sampling time (N/A – not applicable), biological sex (M, male; F, female; U, not recorded), patient age at time of sampling (U, not recorded), and serum IgG, IgA, and IgM concentration.

| Code | Disease | Days | Sex | Age range | Serum Ig conc (mg/mL) |  |  |
| --- | --- | --- | --- | --- | --- | --- | --- |
|  |  |  |  |  | IgG | IgA | IgM |
| 203-0041-1 | Mild | 19 | M | 76-80 | 32.16 | 47.21 | 1.86 |
| 203-0077-1 | Mild | 25 | M | 61-65 | 3.26 | 6.07 | 0.09 |
| 203-0082 | Mild | 21 | F | 71-75 | 20.80 | 15.46 | 2.24 |
| 203-0087 | Mild | 9 | M | 81-85 | 1.94 | 6.50 | 0.24 |
| 203-0091-1 | Mild | 12 | F | 66-70 | 0.58 | 1.69 | 0.01 |
| 203-0112 | Mild | 16 | F | 36-40 | 48.00 | 33.25 | 2.94 |
| 203-0113 | Mild | 8 | M | 66-70 | 59.78 | 63.56 | 5.06 |
| 203-0116 | Mild | 5 | M | 56-60 | 29.93 | 11.86 | 0.82 |
| 203-0009-1 | Moderate | 18 | M | 71-75 | 11.69 | 8.57 | 0.45 |
| 203-0054-1 | Moderate | 9 | M | 46-50 | 13.26 | 4.07 | 0.31 |
| 203-0071 | Moderate | 13 | F | 61-65 | 6.33 | 4.02 | 0.28 |
| 203-0111 | Moderate | 10 | F | 46-50 | 23.01 | 14.09 | 6.11 |
| 203-0132 | Moderate | 16 | M | 51-55 | 17.49 | 41.85 | 2.24 |
| 203-0135 | Moderate | 5 | F | 66-70 | 7.65 | 3.32 | 0.74 |
| 203-0202 | Moderate | 16 | F | 81-85 | 29.49 | 21.12 | 5.39 |
| 203-0203 | Moderate | 14 | M | 76-80 | 21.98 | 48.37 | 2.08 |
| 203-0004-1 | Severe | 18 | M | 41-45 | 51.30 | 25.68 | 2.78 |
| 203-0014 | Severe | 22 | M | 66-70 | 3.10 | 3.57 | 0.15 |
| 203-0015-2 | Severe | 16 | M | 66-70 | 75.25 | 31.57 | 4.83 |
| 203-0018-1 | Severe | 19 | M | 66-70 | 30.53 | 20.31 | 0.71 |
| 203-0021-1 | Severe | 15 | M | 61-65 | 50.99 | 12.93 | 3.71 |
| 203-0023-1 | Severe | 11 | F | 56-60 | 58.52 | 48.17 | 1.79 |
| 203-0024-1 | Severe | 31 | F | 66-70 | 83.84 | 29.13 | 3.61 |
| 203-0025 | Severe | 22 | M | 71-75 | 5.99 | 5.87 | 0.52 |
| 203-0029-1 | Severe | 11 | M | 41-45 | 3.96 | 0.95 | 0.10 |
| 203-0078-2 | Severe | 9 | F | 31-35 | 23.08 | 8.30 | 1.80 |
| NC1 | None | N/A | U | U | 1.89 | 1.96 | 0.07 |
| NC2 | None | N/A | U | U | 2.72 | 1.93 | 0.13 |
| NC3 | None | N/A | U | U | 3.70 | 3.43 | 0.19 |
| NC4 | None | N/A | U | U | 1.98 | 2.45 | 0.19 |
| NC5 | None | N/A | U | U | 2.82 | 3.08 | 0.15 |
| NC6 | None | N/A | U | U | 2.27 | 3.54 | 0.19 |
| NC7 | None | N/A | U | U | 1.81 | 2.43 | 0.21 |
| NC8 | None | N/A | U | U | 3.06 | 3.37 | 0.28 |
| NC9 | None | N/A | U | U | 1.35 | 1.29 | 0.08 |
| NC10 | None | N/A | U | U | 1.79 | 2.18 | 0.10 |

### 2.4. Serum Ig isotype quantification

The IgG, IgA and IgM isotype content in each serum samples was quantified using IgG (total), IgM (total), and IgA (total) human uncoated ELISA kits according to manufacturer‘s instructions. Pre-cleared serum was diluted 1 in 10,000 in phosphate-buffered saline, pH 7.4 (PBS) supplemented with 0.05% (v/v) Tween® 20 (PBS-T) and 0.5% (w/v) bovine serum albumin (BSA) (PBS-T/BSA) for IgA and IgM, and 1 in 500,000 in PBS-T/BSA for IgG quantification. Standard curves were generated in GraphPad Prism (v9.2.0, GraphPad Software, San Diego, CA, U.S.A.) using the sigmoidal, four parameter curve following log transformation of concentration.

### 2.5. Serum antibody fractionation

All serum samples were pre-cleared by centrifugation (10,000 x *g*, 15 min, 4 °C) and 90 μL serum was diluted to a final volume of 400 μL in PBS, pH 7.4, for immediate sequential antibody fractionation (Fig. 1F). Diluted serum (400 μL) was loaded on to a Nab™ protein G spin column and incubated with rotation (60 rpm) at room temperature for 15 min. The protein G column was washed and serum IgG eluted as per manufacturer’s instructions. Elution aliquots with a 280 nm absorbance of >0.01 were pooled, concentrated and buffer-exchanged four times with PBS, pH 7.4, in a 0.5 mL 100 kDa MWCO centrifugal filter. The IgG-depleted protein G column washes containing the unbound material with absorbance of >0.01 at 280 nm were pooled and concentrated in a 0.5 mL 30 kDa MWCO centrifugal filter. The concentrated IgG-depleted protein G column wash (80 μL) was loaded on to a Peptide M agarose column containing 400 μL slurry (200 uL matrix) and incubated with rotation (60 rpm) at room temperature for 15 min. The Peptide M column was washed and serum IgA was eluted as per manufacturer’s instructions. Eluate with a 280 nm absorbance of >0.01 was pooled, concentrated and buffer-exchanged with PBS, pH 7.4, as above. IgG- and IgA-depleted column washes with absorbance of >0.01 at 280 nm were pooled and concentrated in a 0.5 mL 30 kDa MWCO centrifugal filter and (80 μL) loaded on to a POROS CaptureSelect™ affinity column containing 400 μL slurry (200 uL matrix). The loaded column was incubated with rotation (60 rpm) at room temperature for 15 min, and the column was washed and serum IgM was eluted according to manufacturer’s instructions. Wash and elution fractions of absorbance >0.01 at 280 nm were pooled separately, and eluate was concentrated and buffer-exchanged with PBS, pH 7.4, in a 0.5 mL 100 kDa MWCO centrifugal filter as above.

**Fig. 1.**
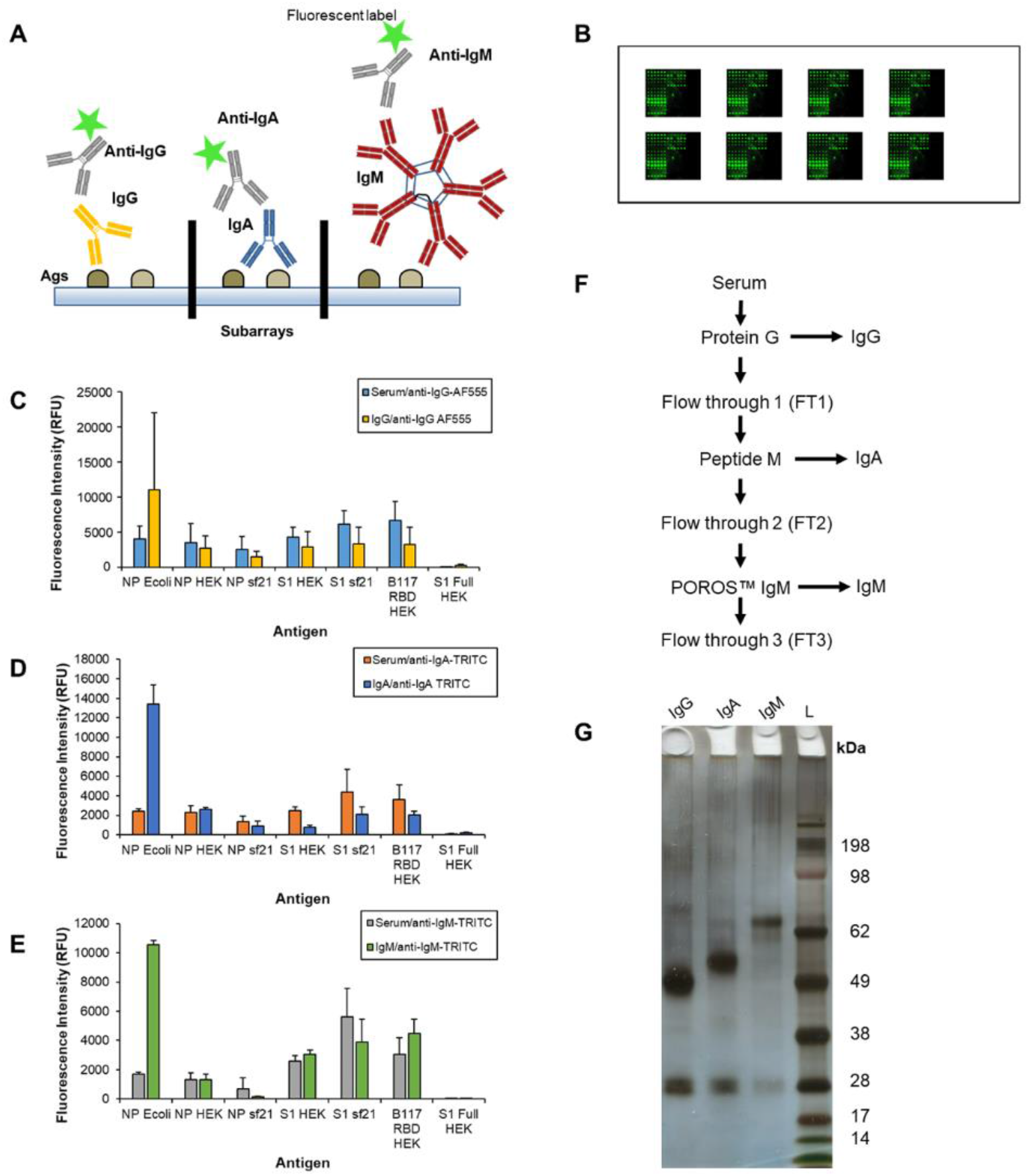
Construction and optimisation of a SARS-CoV-2 protein antigen microarray and fractionation of serum immunoglobulins. (A) Cartoon of SARS-CoV-2 protein antigens conjugated to a microarray surface in replicate subarray format. Each subarray was incubated with a purified serum antibody isotype and serum antibody binding to specific antigens was detected using fluorescently labelled anti-isotype antibodies. (B) Cartoon of a typical SARS-CoV-2 antigen microarray slide with six replicate subarrays. (C) Bar charts representing the binding intensity of diluted serum (1/100 in PBS-T) and purified serum (C) IgG, (D) IgA, and (E) IgM from a COVID-19 patient (203-0018-1) detected by the appropriate fluorescently-labelled anti-isotype antibody. (F) Flow chart depicting sequential serum immunoglobulin isotype purification procedure. (G) Purified serum IgG, IgA, and IgM (1 μg each) from a COVID-19 patient (203-0025) electrophoresed on a 4-12% Bis-Tris SDS-PAGE gel and silver stained. L, molecular mass ladder (kDa).

All recovered purified serum IgG, IgA, and IgM were quantified by BCA assay using BSA as the standard. Antibody isotype recovery was verified by SDS-PAGE gel electrophoresis on a 4-12% Bis-Tris gel (1 μg of each antibody loaded) under reducing conditions with MOPS running buffer and 0.05% Coomassie G-250 staining [36] (Fig. 1G). Serum antibody samples were aliquoted and stored at -20 °C until further use.

### 2.6. SARS-CoV-2 protein antigen microarray construction

Just before printing, the protein antigens Npro Full Ecoli, Spro Ecoli, S1Frag Ecoli, RBD Ecoli, S2Frag Ecoli, S2Pri Ecoli, and MP Ecoli were buffer exchanged with PBS, pH 7.4, and quantified using absorbance at 280 nm on a NanoDrop™ 2000/2000c spectrophotometer and a BSA standard curve. All protein antigens in PBS, pH 7.4 (Table 1) were printed on Nexterion® H microarray slides in six replicate features, approximately 1 nL per feature (2 drops), with 8 replicate subarrays per microarray slide using a Scienion SciFlexArrayer S3 essentially as previously described [37]. Slides were incubated overnight at 19 °C in a high humidity chamber to complete conjugation. Microarrays were incubated in 100 mM ethanolamine in 50 mM sodium borate, pH 8.0, for 1 h at room temperature to deactivate remaining functional groups. Slides were washed three times in PBS-T for 5 min each wash, then once in PBS. Finally, microarrays were centrifuged dry (475 x *g*, 5 min, 15 °C) and stored at 4 °C with desiccant. Antigen microarrays were used within 1 month of construction.

### 2.7. Profiling serum antibody binding to SARS-CoV-2 protein antigens

Serum antibody profiling was carried out in a two-step incubation process. Serum IgG and IgA were diluted to 10 μg/mL in PBS-T and serum IgM diluted to 8-12 μg/mL in PBS-T. Seventy μL of each antibody dilution was incubated in a separate antigen subarray for 1 h at 23 °C with gentle inversion (4 rpm) as previously described [37]. For the first microarray incubation step, serum IgG, IgA and IgM from two individuals (six subarrays) were incubated and the two remaining subarrays were incubated with PBS-T. Slides were then washed twice in PBS-T and once in PBS prior to centrifuging dry as above. For the second step, 70 μL of fluorescently labelled anti-human IgG, IgA, and IgM diluted in PBS-T (1 in 5,000, 1 in 1,000 and 1 in 1,000, respectively), were applied to the corresponding subarrays and in the two previously ‘blank’ subarrays a fluorescently labelled lectin and a secondary antibody control were incubated per antigen microarray. A selection of TRITC-labelled lectins were added at 10 or 15 μg/mL diluted in Tris-buffered saline (TBS; 20 mM Tris-HCl, 100 mM NaCl, 1 mM CaCl_2_, MgCl_2_, pH 7.2) with 0.05% Tween-20 (TBS-T) (Table S2). For the second step, slides were again incubated for 1 h at 23 °C with gentle inversion (4 rpm), then washed twice in PBS-T, once in PBS, and centrifuged dry. Slides were scanned immediately in an Agilent G2505 microarray scanner (Agilent Technologies, Cork, Ireland) (532 nm laser, 90% PMT, 5 μm resolution) and images saved as .tif files.

### 2.8. Microarray data extraction and analysis

Data extraction was performed essentially as previously described [37]. In brief, raw intensity values were extracted from the image *.tif files using GenePix Pro v6.1.0.4 (Molecular Devices, Berkshire, UK) and a proprietary *.gal file (containing feature spot address and identity) using adaptive diameter (70–130%) circular alignment based on 230 μm features and were exported as text to Excel (Version 2016, Microsoft). Local background-corrected median feature intensity data (F532median-B532) was analysed. The median of six replicate spots per subarray was handled as a single data point for graphical and statistical analysis and considered as one experiment. Data were normalised to the per-subarray mean total intensity value of the three technical replicate microarray slides (or duplicate in specified cases). Binding data was presented in bar chart form of the average intensity of three experimental replicates +/- one standard deviation (SD).

### 2.9. Statistical analysis of serum antibody binding to antigens

For each of IgG, IgA and IgM the antigen binding intensity was compared using a separate linear mixed model (LMM). The LMM framework is useful to account for the correlation of the outcome within an individual’s replicates. The 13 antigens were split into those seven which were high binding (NP Ecoli, NP HEK, NP Sf21, S1 HEK, S1 Sf21, B117 RBD HEK, S1 Full HEK) and six which were low binding (S2Frag Ecoli, S2Pri Ecoli, S1Frag Ecoli, Npro Full Ecoli, Spro Ecoli, 3CLike Ecoli), resulting in six total models. In each of the six LMM the COVID-19 severity (none, mild, moderate, severe) and antigen was included. An interaction term between COVID-19 severity and antigen was added to investigate whether certain antigens had higher binding intensity in those with more severe disease. This interaction term was the key estimate – the difference in binding intensity across COVID-19 severity is reported for each of the 13 antigens along with 95% confidence intervals. With 13 antigens, significance is taken to be p<0.0038 (Bonferroni adjusted 0.05/13).

## 3. Results

### 3.1. Serum collection and antibody isotype quantification and purification

Serum from 26 patients admitted to hospital with COVID-19 symptoms and confirmed by RT-qPCR as SARS-CoV-2 infected were obtained and categorised by COVID-19 disease severity as mild, moderate or severe according to WHO scale classification [38, 39] (Table 2). Mild was defined as COVID-19 patients who were managed on the ward and did not require ICU admission or escalation, and excluded patients who were palliated (WHO scores 4 to 5). Moderate defined COVID-19 patients who were managed with high flow or non-invasive ventilation either in the ICU or on the ward (WHO score 6). Patients with severe disease were those who required invasive mechanical ventilation in the ICU and/or patients who died from the disease (WHO scores 7 to 10). Additionally, 10 serum samples from healthy individuals who donated blood prior to November 2018 (i.e. pre-COVID-19 pandemic) were used as ‘non-COVID-19’ (NC) controls. The total IgG, IgA and IgM concentration was quantified from sera using dedicated ELISAs (Table 2) and IgG, IgA, and IgM were purified from each serum sample by sequential affinity chromatography (Fig. 1F and G).

### 3.2. SARS-CoV-2 antigen microarray construction and optimisation

Several presentation strategies were investigated in parallel to assess for sensitive and robust detection of serum antibody isotype binding to specific recombinant SARS-CoV-2 protein antigens. Initially IgG, IgA, and IgM purified from a SARS-CoV-2 positive serum sample along with the IgG-, IgM- and IgA-depleted flow through were printed on a microarray to assess the specificity of fluorescently labelled anti-isotype secondary antibodies. Cross-reactivity from anti-IgG with IgA and IgM was noted, whereas anti-IgA and anti-IgM were specific for their respective isotype (Fig. S1). Thus, the use of purified serum isotypes was continued.

Next, antigen binding to the antibody isotypes purified from a SARS-CoV-2 positive serum sample printed on microarrays was assessed. The binding of NP Ecoli, 3CLike Ecoli, and B117 RBD HEK (Table 1) to immobilised antibody isotypes was detected using a fluorescently labelled anti-His antibody (Fig. S2). No binding from 3CLike Ecoli and B117 RBD HEK was noted, but NP Ecoli bound to the immobilised serum antibody isotypes in a concentration dependent manner. However, binding intensities achieved were too low (approximately 0-700 RFU) to feasibly use a printed serum antibody isotype microarray to detect low levels of antigen binding and this approach was not pursued.

An alternative strategy of printing a panel of immunologically relevant SARS-CoV-2 recombinant protein antigens, focussing on NP and S proteins, on microarray slides to create a SARS-CoV-2 antigen microarray (Table 1) was also pursued. Binding of serum antibodies to the antigens was initially evaluated using diluted serum and detected using fluorescently-labelled anti-isotype antibodies incubated in separate subarrays (Fig. S3). Overall serum antibody binding intensities to antigens were increased compared to the immobilised serum antibody isotype format (to approximately 0-10,000 RFU) (Fig. S3) and were further improved using purified serum antibody isotypes (Figs. 1C, D, E and S4), which also removed any cross-reactivity issues from anti-isotype antibodies or potential competitive binding between serum antibody isotypes. It is worth noting that the binding pattern of the purified antibodies were altered compared to the complex serum sample.

### 3.3. Disease severity discrimination using serum antibody isotype binding to SARS-CoV-2 antigens

Purified serum antibody isotypes from non-COVID-19 donors (NC) and COVID-19 patients with mild, moderate, and severe disease (Table 2) were incubated on printed antigen microarrays. Antibody isotypes were detected in separate subarrays using their respective fluorescently labelled anti-isotype antibody (Fig. 2 and Table S3). The C-terminal membrane glycoprotein (MP Ecoli) antigen did not conjugate to the microarray surface and is omitted from presented data. A range of binding intensities across the detection range of the microarray scanner (approximately 100-65,500 RFU) were observed, demonstrating sensitive and robust detection. Immune recognition of the antigens varied by individual COVID-19 patient, with not all patients recognising the same selection of antigens, and the intensity of antibody binding generally varied by disease severity. In general, for the serum antibody isotype binding to the antigens, the binding intensity of IgG was greatest, followed by IgA, with IgM exhibiting lowest intensity binding.

**Fig. 2.**
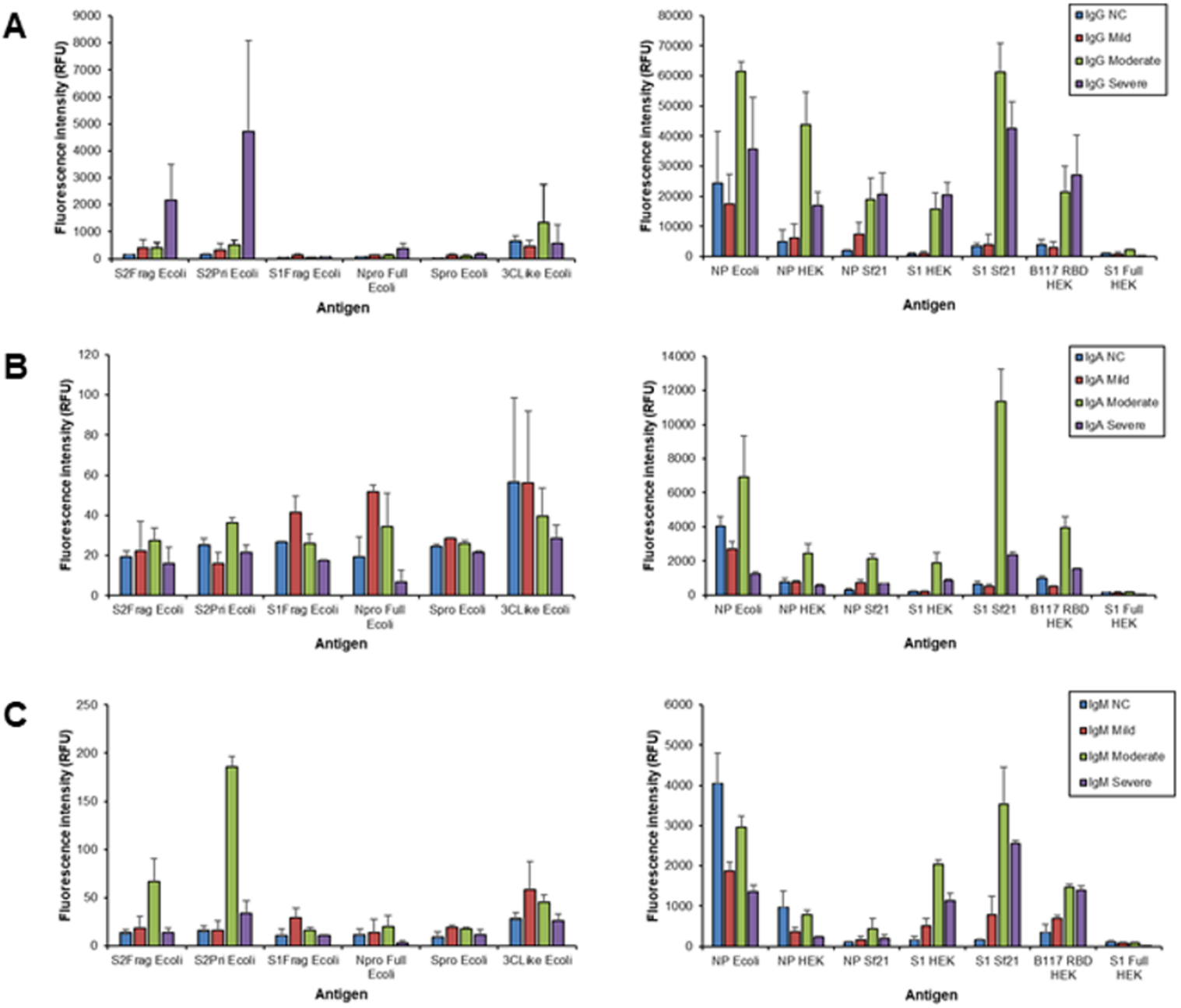
Bar charts representing the binding intensity of purified antibody isotypes from a selected patient from each cohort to immobilised antigen detected by fluorescently-labelled anti-isotype antibodies. (A) IgG, (B) IgA, and (C) IgM. Non-COVID-19 (NC) sample NC1, mild patient 203-0077, moderate 203-0009, and severe 203-0004. Data from the same experiments are represented as two separate bar charts, one for low and one for high binding intensities.

For serum IgG there was a significant association between COVID-19 disease severity and binding intensity for the Npro Full Ecoli, S1Frag Ecoli, Spro Ecoli, S2Pri Ecoli, S2Frag, NP Ecoli, NP HEK and S1 Sf21 antigens (Fig. 3, Tables S4 and S5). In particular, there was a significant average decrease in intensity for the Npro Full Ecoli antigen among those with moderate disease compared to those without COVID-19. S1Frag Ecoli and Spro Ecoli binding was higher on average in those with mild COVID-19 compared to those without. There was an average increase in binding to both the S2Pri Ecoli and S2Frag Ecoli antigens in those with mild/severe COVID-19, compared to those without. However, those with moderate COVID-19 had lower binding to S2Pri Ecoli on average compared to those without COVID-19. There was also a significant average decrease in IgG binding intensity for the NP Ecoli antigen among those with mild and moderate disease compared to those without COVID-19. There was an average increase in IgG binding to both the NP HEK and S1 Sf21 antigens in those with severe COVID-19, compared to those without.

**Fig. 3.**
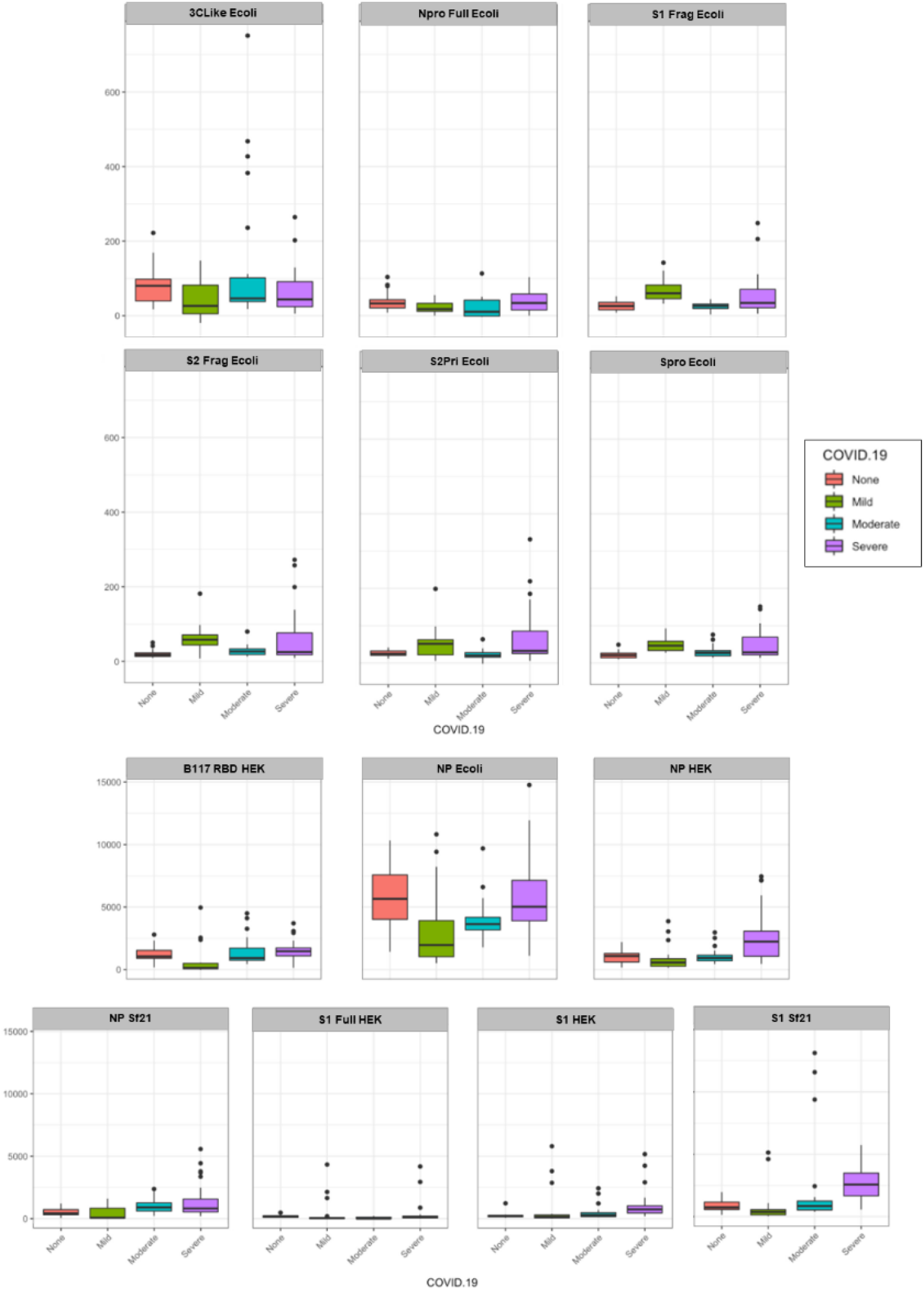
Serum IgG samples by COVID-19 disease severity (mild, moderate, severe) binding to the various SARS-CoV-2 protein antigens compared to no disease (NC). Each antigen was plotted using a boxplot with relative fluorescence intensity of binding on the y-axis and COVID-19 disease cohort on the x-axis.

For serum IgA, there was a significant association between COVID-19 severity and binding intensity for the Npro Full Ecoli, S1Frag Ecoli, S2Frag Ecoli, S2Pri Ecoli, Spro Ecoli, S1 Full HEK and S1 Sf21 (Fig. 4, Tables S6 and S7). In particular, there was a significant average decrease in intensity for the NPro Full Ecoli, S1Frag Ecoli, S2Frag Ecoli, S2Pri Ecoli and Spro Ecoli antigens among those with moderate disease compared to those without COVID-19. There was also a significant average decrease in binding intensity for the S1 Full HEK and increased average S1 Sf21 binding among those with severe disease compared to those without COVID-19.

**Fig. 4.**
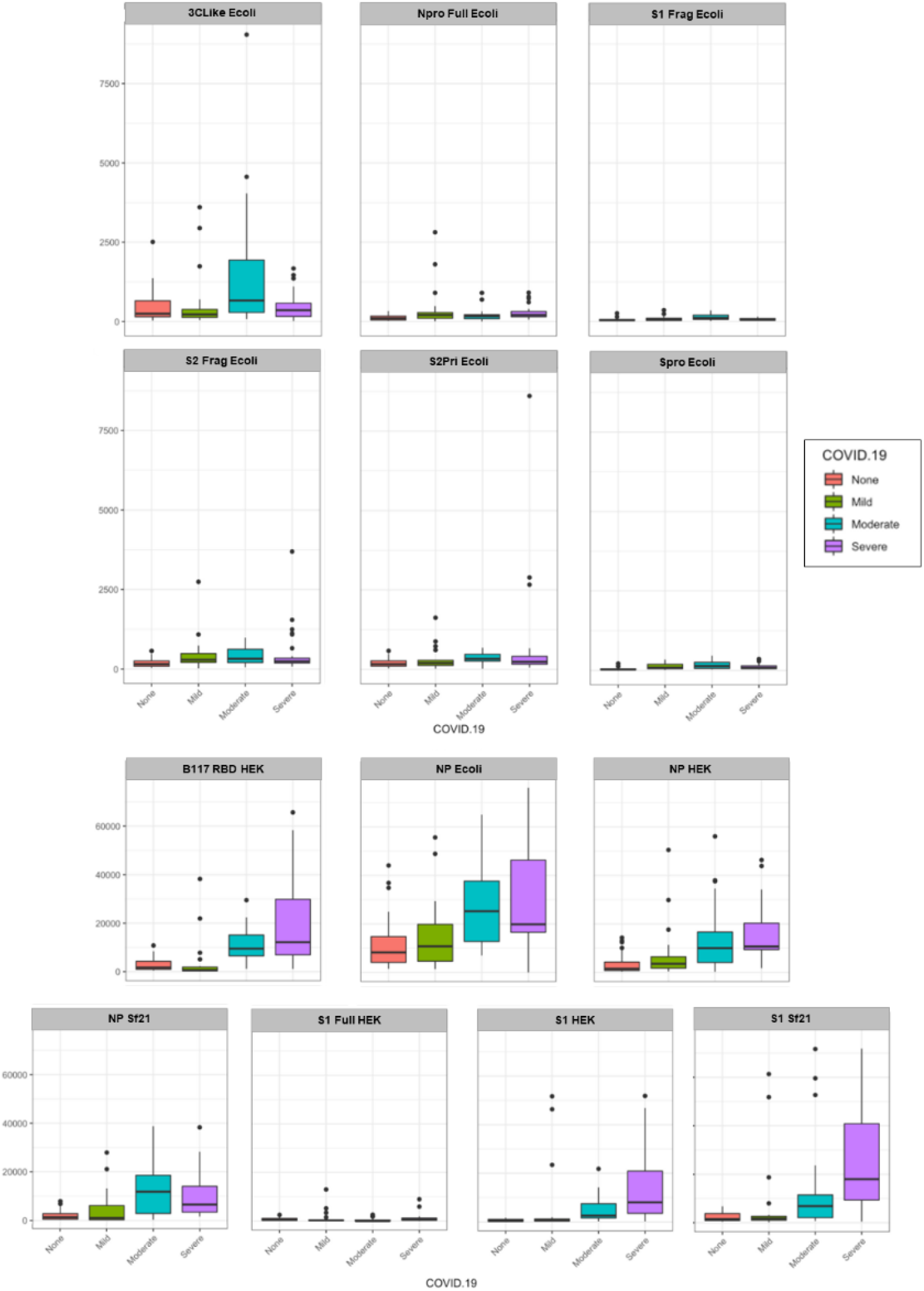
Serum IgA samples by COVID-19 disease severity (mild, moderate, severe) binding to the various SARS-CoV-2 protein antigens compared to no disease (NC). Each antigen was plotted using a boxplot with relative fluorescence intensity of binding on the y-axis and COVID-19 disease cohort on the x-axis.

Serum IgM demonstrated a significant association between COVID-19 severity and binding intensity for the Npro Full Ecoli, S2Frag Ecoli and S2Pri Ecoli, S1 Full HEK, and S1 Sf21 antigens (Fig. 5, Tables S8 and S9). In particular, there was a significant average decrease in intensity for the Npro Full Ecoli, S2Frag Ecoli and S2Pri Ecoli antigens among those with mild disease compared to those without COVID-19, and a significant decrease in average intensity for the S1 Full HEK and increase in average S1 Sf21 binding among those with severe disease compared to those without COVID-19.

**Fig. 5.**
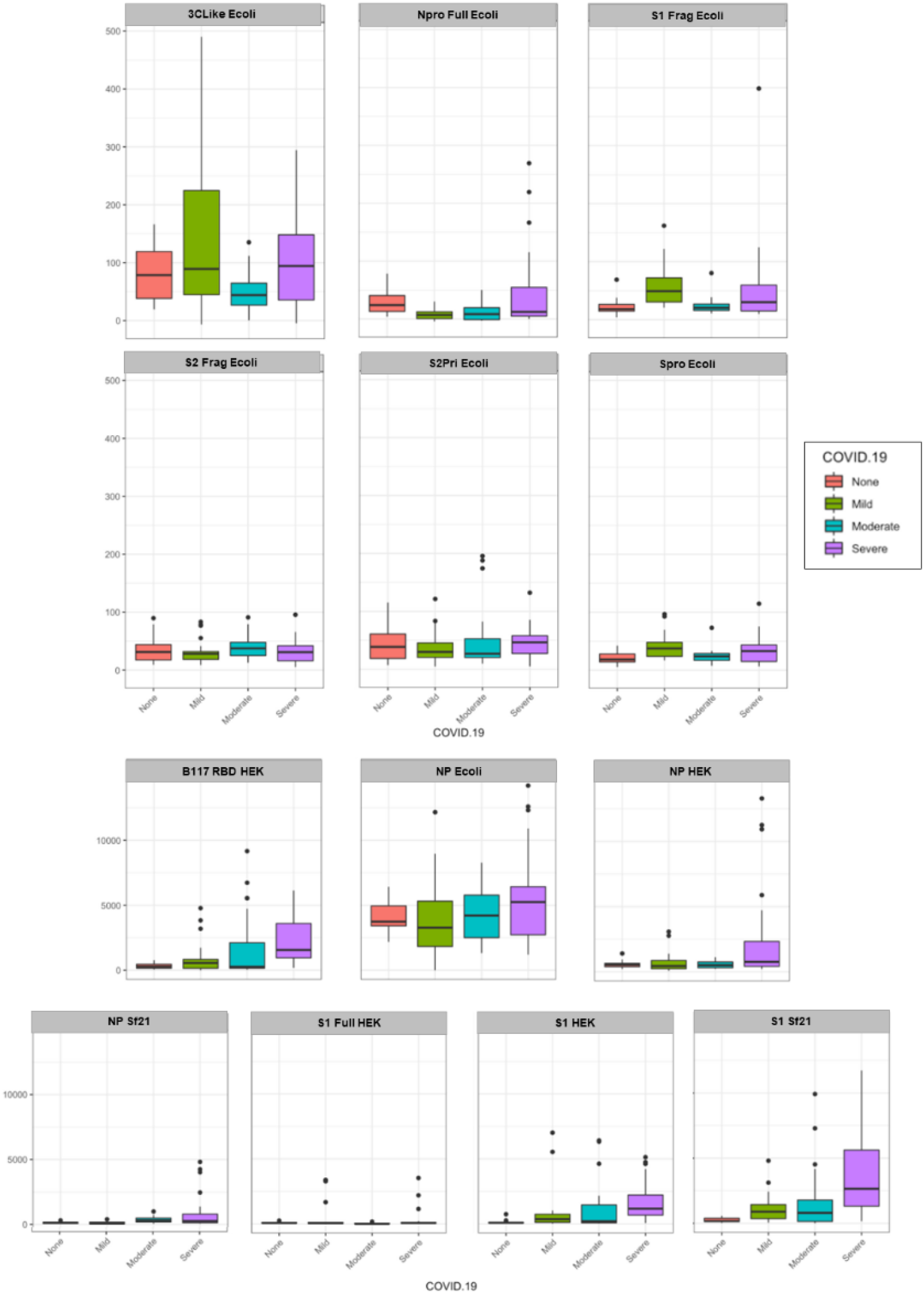
Serum IgM samples by COVID-19 disease severity (mild, moderate, severe) binding to the various SARS-CoV-2 protein antigens compared to no disease (NC). Each antigen was plotted using a boxplot with relative fluorescence intensity of binding on the y-axis and COVID-19 disease cohort on the x-axis.

### 3.4. Antigen glycosylation influences serum antibody recognition

Proteins recombinantly expressed in *E. coli* are missing post-translational modifications (PTMs) while those expressed in mammalian and insect cells can have PTMs, including glycosylation, but the PTMs and glycosylation will vary depending on the system used with consequent impacts on protein functionality [40]. In human infections, the SARS-CoV-2 S protein is very heavily glycosylated with both high mannose and complex type N-linked glycosylation and O-linked glycosylation, which shields much of the viral protein backbone from host immune recognition including the enfolded RBD [41, 42]. Coronavirus NP on the other hand was previously reported to be mainly phosphorylated [43] but, depending on the sequence and expression system, N- and O-linked glycosylation and phosphorylation of SARS-CoV-2 NP has been reported [44]. As the amount and type of glycosylation can affect antibody and receptor recognition, the use of a correctly modified viral protein antigen is key to a realistic assessment of immune interactions for serological or diagnostic assays.

The S1 Full HEK and S1 HEK antigens demonstrated complex type N-linked glycosylation with α-(2,3)- and α-(2,6)-linked sialylation (MAA and SNA-I binding, respectively) and prominent terminal *N*-acetylgactosamine (GalNAc) residues (WFA) when profiled with lectins, with S1 Full HEK displaying slightly less α-(2,3)-sialylation compared to S1 HEK (Fig. S5). The B117 RBD HEK demonstrated a similar glycosylation profile to S1 Full HEK, with slightly lower terminal GalNAc residues and α-(2,3)-sialylation. S1 Sf21 on the other hand exhibited a profile indicative of high mannose structures (intense HHA, Con A, and WGA binding) (Fig. S5). NP HEK and NK Sf21 both expressed complex type glycosylation, but there was substantially less terminal GalNAc residues and sialylation in NP Sf21 compared to NP HEK (lower MAA, SNA-I, WFA, and SJA binding) while UEA-I additionally bound to NP HEK which indicated fucosylation (Fig. S5).

Overall, there was substantially lower quantitative serum antibody binding to the *E. coli*-produced antigens compared to HEK and Sf21 cell-produced antigens, except in the case of NP Ecoli (Figs. 2-5). Despite this, several *E. coli*-produced antigens remained significant for discriminating disease severity, including Npro Full Ecoli and S2Frag Ecoli which were previously reported to improve SARS-CoV-2 diagnostic accuracy when used in a dual ELISA format [21]. Comparing the serum antibody recognition of NP Ecoli, NP HEK, and NP Sf21, NP glycosylation decreased binding intensity for serum IgG, IgA, and IgM for all disease severity and NC cohorts (Figs. 2-5). Differences in binding to NP HEK remained significant for IgG in severe disease (Table S5) and IgM for moderate disease (Table S9), to NP Sf21 for IgA and IgM binding in severe disease (Table S7 and S9), and IgM for moderate disease (Table S9). The binding of serum IgG, IgA and IgM to S1 Sf21 was higher for all cohorts compared to S1 HEK, with binding to both S1 Sf21 and S1 HEK both substantially higher compared to the most similar *E. coli*-produced protein sequence S1Frag Ecoli (Figs. 2-5). Differences in binding to S1 HEK remained significant for IgG in mild disease, and to S1 Sf21 for IgG binding in moderate disease, and IgG, IgA, and IgM in severe disease (Table S5 and S7).

### 3.5. Variation in antibody binding to viral antigens over time

Ten patient sera were sampled at a later time point in convalescence, two patients with mild, one with moderate and seven with severe disease (Tables S10 and S3). The same concentration of purified serum antibody isotype was incubated on the antigen microarray for all samples to enable comparison of relative proportion of virus-specific antibodies that remained or developed over time. The absolute concentration of antibody isotypes in serum varied over time, generally decreasing by 1 month PFSO (Table S10). However, serum antibody isotype binding intensity to the various antigens changed over time to differing degrees, some decreasing, staying the same or increasing depending on antibody isotype, duration PFSO, and patient disease severity (Figs. 6-8, S6, and S7). Generally for this small cohort of patient samples, antibody isotype binding for mild patients to antigens reached their maximum antigen binding earlier PFSO while those of the severe cohort reached their maximum later.

**Fig. 6.**
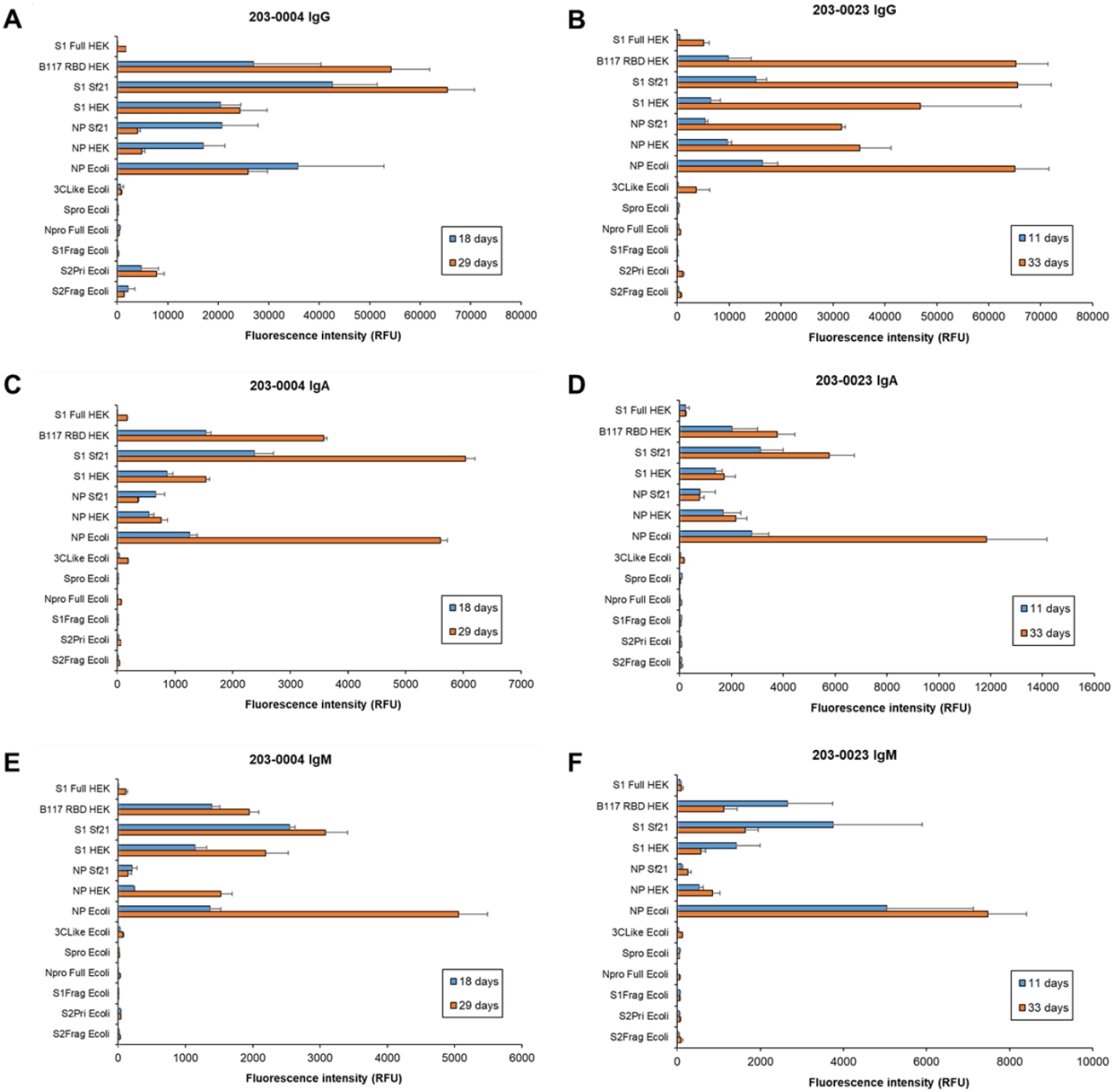
Dynamic binding of patient serum antibody isotypes over short to medium term for severe COVID-19 disease. Bar charts represent binding intensity data for serum (A,B) IgG, (C,D) IgA, and (E,F) IgM binding to SARS-CoV-2 antigens for patient 203-0004 (**A**,**C**,**D**; M, 41-45 years) and 203-0023 (**B**,**D**,**F**; F, 55-60 years) at 18 and 29 days, and 11 and 33 days post-first symptom onset, respectively. Bars represent the average binding intensity from three replicate experiments with error bars of +/- one standard deviation (SD).

For the severe patient sera up to approximately 1 month PFSO, overall antibody binding intensities to the recognised antigens typically increased from the earlier sampling time points (Figs. 6 and S6B,D and F), despite a general decrease in absolute antibody concentrations in serum over time (Table S10). However, variations in the relative binding to various antigens were apparent for some individuals over time. For example, the relative proportion of patient 203-0004 serum IgGs binding to the NP antigens NP Ecoli, NP Sf21, and NP HEK decreased from 18-29 days but increased for the S antigens S1 Full HEK, B117 RBD HEK, S1 Sf21, S1 HEK, and S2Pri Ecoli (Fig. 6A). That of patient 203-0023 serum IgMs binding to the NP antigens NP Ecoli, NP Sf21, and NP HEK increased from 11 to 29 days PFSO but decreased for the S antigens S1 Full HEK, B117 RBD HEK, S1 Sf21, and S1 HEK (Fig. 6F). Similar substantial and varied binding alterations were evident over shorter time intervals for other individual severe patient sera (days 15 and 17, Fig. S6A,C,E, and days 31 and 34, Fig. S7A,C,E).

The response of serum IgG, IgA, and IgM for moderate disease patient 203-0054 from 9 to 12 days PFSO displayed a substantial increase in binding to antigens over the 3 day interval (Fig. 7A,C, and E). Notably for this moderate disease patient, IgA and IgM binding intensities were similar in magnitude to IgG. IgG and IgA binding intensities for mild disease (patients 203-0041 and 203-0077) generally were greatest at the earlier time points (19 and 25 days PFSO, respectively) and decreased over medium term (by 34 and 59 days PFSO, respectively) (Figs. 7B,D,F and S7B,D,F). However, for mild disease serum IgM, binding intensity alterations over time were mixed depending on the individual, and actually increased in response to certain antigens (e.g. increase in 203-0041 IgM binding to S1 Sf21 by 34 days PFSO and 203-0077 IgM binding to NP E coli) (Figs. 7F and S7F).

**Fig. 7.**
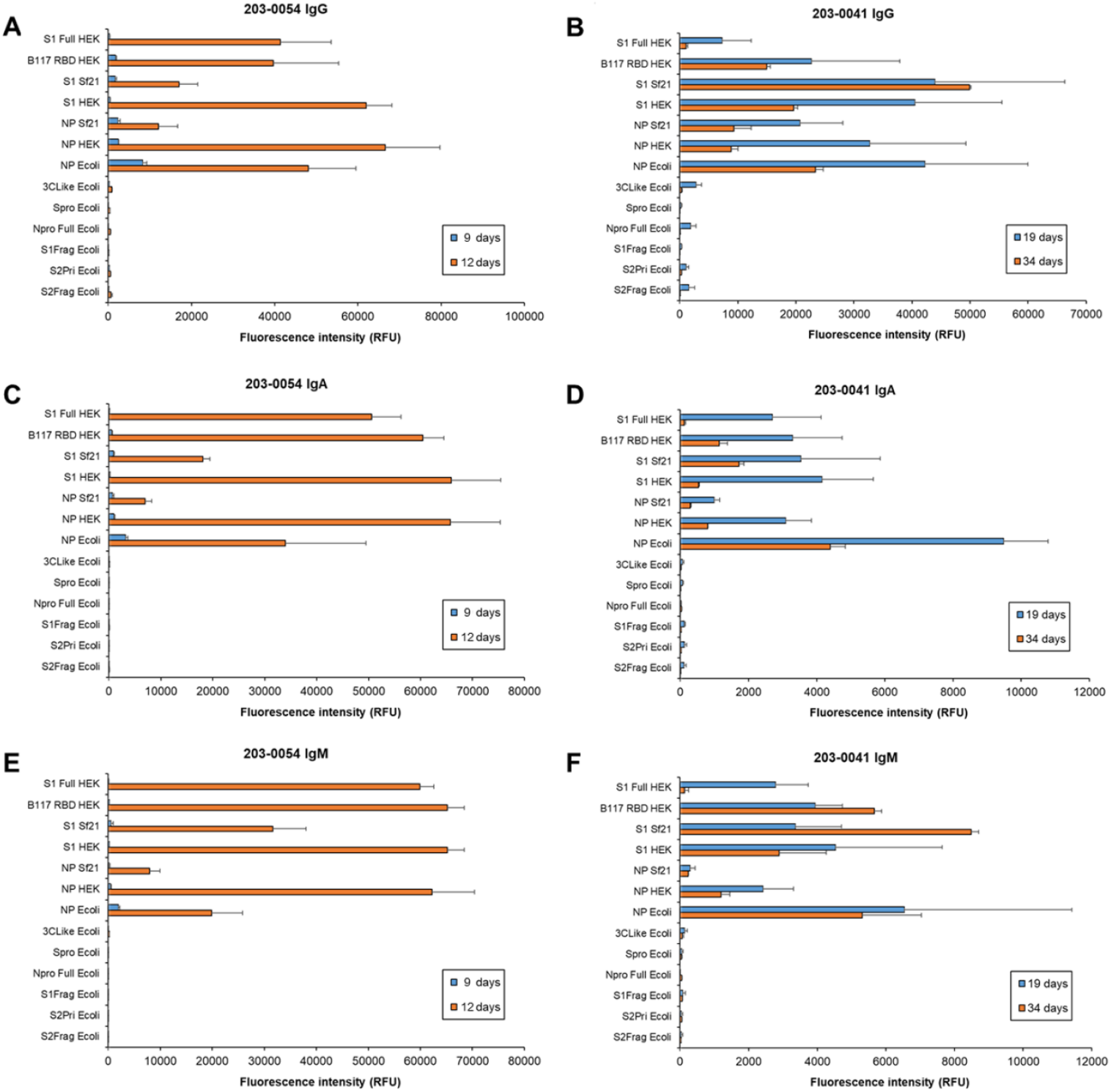
Dynamic binding of patient serum antibody isotypes over short to medium term for moderate and mild COVID-19 disease. Bar charts represent binding intensity data for serum (A,B) IgG, (C,D) IgA, and (E,F) IgM binding to SARS-CoV-2 antigens for patient 203-0054 (**A**,**C**,**D**; moderate, M, 46-50 years) and 203-0041 (**B**,**D**,**F**; mild, M, 76-80 years) at 9 and 12 days, and 19 and 34 days post-first symptom onset, respectively. Bars represent the average binding intensity from three replicate experiments with error bars of +/-1 SD.

For two severe patient samples 203-0015 and 203-0018 with longer-term follow up samples (145 and 271 days PFSO, respectively), IgG demonstrated highest binding intensity to SARS-CoV-2 antigens at the earlier time PFSO, 16 and 19 days respectively, and the binding to all antigens substantially decreased by the approximately 5 and 9 months PFSO (Fig. 8A and B), respectively. However, IgG binding was still detected at the late convalescence timepoints using this sensitive platform.

**Fig. 8.**
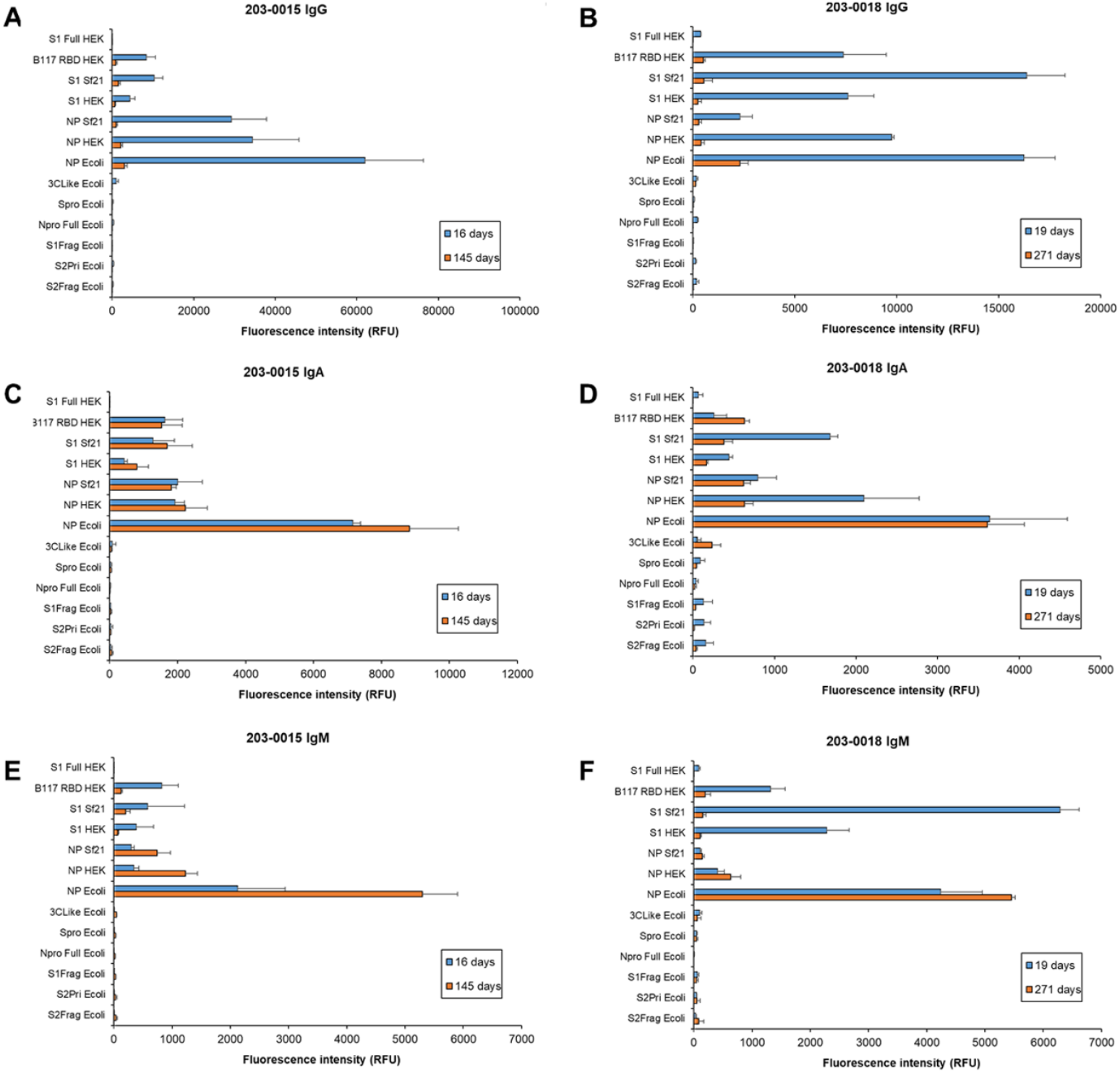
Dynamic binding of patient serum antibody isotypes over longer term for severe COVID-19 disease. Bar charts represent binding intensity data for serum (A,B) IgG, (C,D) IgA, and (E,F) IgM binding to SARS-CoV-2 antigens for patient 203-0015 (**A**,**C**,**D**; M, 66-70 years) and 203-0018 (**B**,**D**,**F**; M, 66-70 years) at 16 and 145 days, and 19 and 271 days post-first symptom onset, respectively. Bars represent the average binding intensity from three replicate experiments with error bars of +/- 1 SD.

On the other hand, IgA intensity for the recognised antigens remained at a similar binding intensity level after 5 months for patient 203-0015, and either increased or remained the same for 3CLike Ecoli, NP Ecoli, and B117 RBD HEK and decreased for the other antigens initially bound for patient 203-0018 9 months PFSO (Fig. 8C and D). Patient 203-0015 serum IgM binding increased for the NP antigens NP Ecoli, NP HEK and NP Sf21 but decreased for the S protein antigens B117 RBD HEK, S1 Sf21, and S1 HEK by 5 months (Fig. 8E). The 9 month PFSO IgM response was similar for patient 203-0018, with either slightly increased or similar level of response for NP antigens but a significant decrease for the S antigens S1 Full HEK, B117 RBD HEK, S1 Sf21, and S1 HEK (Fig. 8F). It is worth noting that the low intensity binding of 203-0018 IgG, IgA, and IgM to S1 Full HEK was absent by approximately 9 months PFSO, while 203-0015 serum antibodies had not developed a response to the same antigen at 16 or 145 days (Fig. 8).

### 3.6. Post-vaccination serum antibody isotype binding to SARS-CoV-2 antigens

Serum antibody isotypes were purified from two healthy donors who had not been previously infected with SARS-CoV-2 30 days after their second vaccination with the Pfizer-BioNTech COVID-19 mRNA vaccine, which contains nucleoside-modified mRNA encoding the SARS-CoV-2 S glycoprotein [45]. Overall, the donor binding intensities was dissimilar to the diversity of responses of COVID-19 patients to the presented viral antigens (Fig. 9). Donors demonstrated high intensity binding of serum IgG to S proteins B117 RBD HEK, Sf Sf21 and S1 HEK, with donor V2 higher than V1, but not to other presented viral antigens (Fig. 9A), demonstrating specific S protein response as expected due to the presentation of only S protein in the mRNA vaccine but numerous antigens during infection. Both donors also had a proportionally similar response to the antigens (i.e. S1 Sf21 > B117 RBD HEK > S1 HEK). Both donors had low IgG binding intensity to S1 Full HEK, similar to the low binding intensity level to the NP antigens NP Sf21, NP HEK, and NP Ecoli persisting from previous endemic coronavirus exposure of the donor.

**Fig. 9.**
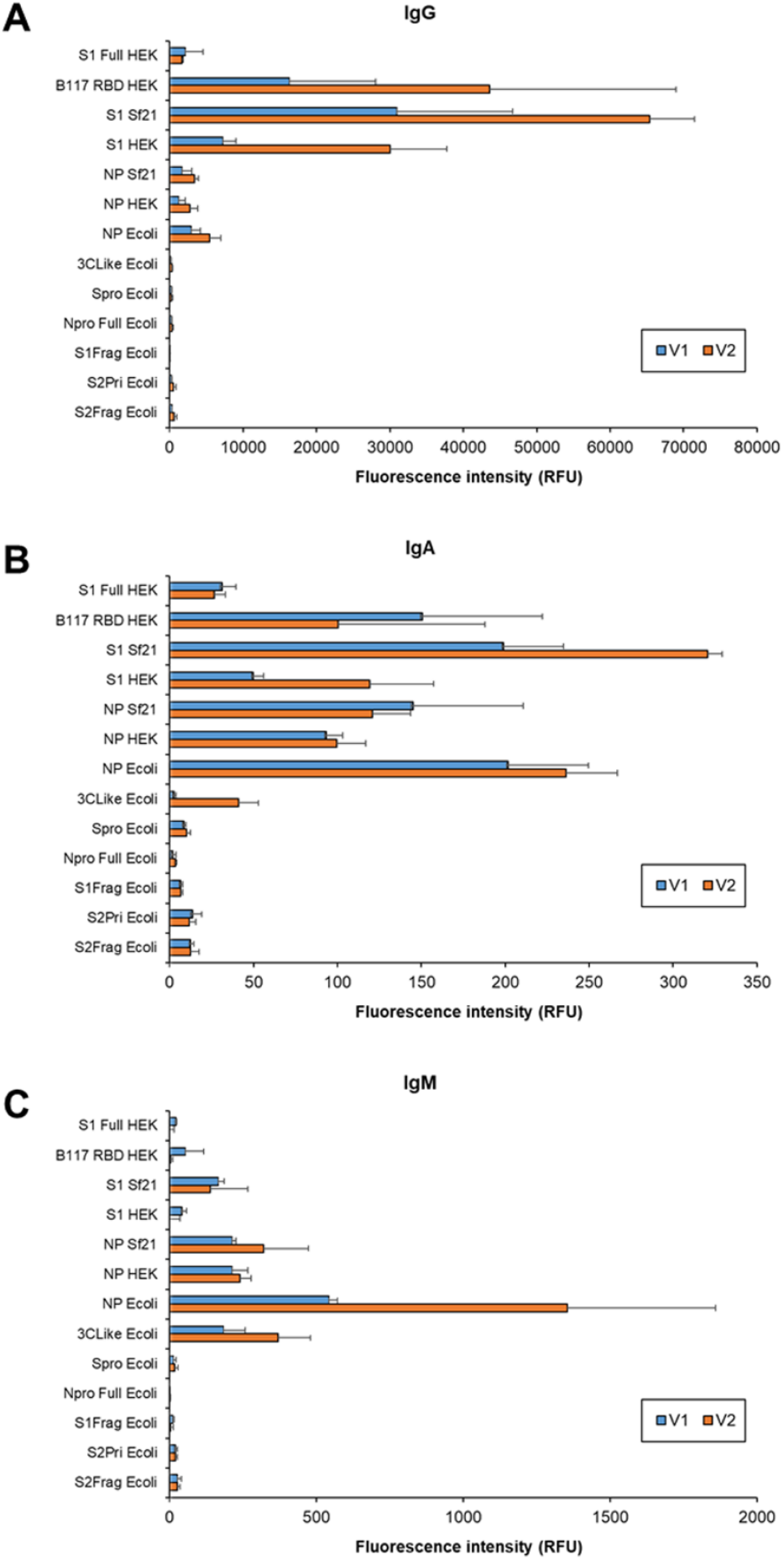
Serum antibody response of double vaccinated healthy donors with no previous SARS-CoV-2 infection. Bar charts represent binding intensity of purified serum (A) IgG, (B) IgA, and (C) IgM from two double vaccinated healthy donors V1 (M, 56-60 years old) and V2 (F, 31-35 years old) with no previous SARS-CoV-2 infection (V1 and V2) to immobilised antigen detected by fluorescently-labelled anti-isotype antibodies. Bars represent the average binding intensity from three replicate experiments with error bars of +/- 1 SD.

The binding intensity of serum IgA and IgM 30 days after the second mRNA vaccine dose remained low to negligible for both vaccinated healthy donors, with binding to S proteins at similar intensity levels as previous exposures to endemic coronavirus NP for IgA (Fig. 9B). On the other hand, IgM binding to S antigens was lower than to endemic coronavirus NP (Fig. 9C), indicating essentially no IgM response to the mRNA vaccine. These data suggest that vaccination with mRNA for S protein or protein fragment did not elicit the same substantial IgA and IgM response as SARS-CoV-2 infection.

## 4. Discussion

In this work, we developed and employed a multiplexed SARS-CoV-2 antigen microarray platform for profiling and quantifying the binding of constant concentrations of purified serum IgG, IgA, and IgM to a panel of antigens simultaneously within the same experiment. We were able to correlate the binding intensities of the antibody isotypes to panels of antigens with COVID-19 disease severity and note the impact of antigen glycosylation on antibody binding. We also observed the persistence of the same relative proportion of all antigen-specific serum IgA binding over longer convalescence periods, while the proportion of antigen-specific IgG declined over time. On the other hand, the relative proportion of IgM binding to S antigens declined over time but were maintained or increased for NP antigens.

Our multiplexed platform was sensitive, accurate, and reproducible without the need for a separate blocking step or inclusion of blocking agent in the applied sample that is required for the majority of other reported multiplexed platforms [26-34]. We observed differences in quantitative and relative binding pattern between purified serum IgG, IgA, and IgM compared to the binding of same isotypes in the presence of all others in serum dilutions detected by anti-isotype secondary antibodies. These differences were likely due to several factors. Firstly, some anti-isotype secondary antibodies can have cross-reactivity with other isotypes, as demonstrated in this work where anti-IgG antibody also bound to IgA and IgM (Fig. S1), which can artefactually increase or decrease signals for individual isotypes. Competition between antibody isotypes for the same antigen can also confound individual isotypes binding data when using a mixture of isotypes in the sample, and can require the inclusion of additional reagents to decrease competition when it is noticed. Dobaño, *et al*. used GullSORB™, a reagent for precipitating IgG, to increase IgM binding signal and reduce background in their multiplexed serological assay using diluted plasma samples [26]. Competition between IgG, IgA, and IgM isotypes for glycan antigen binding has also been previously reported [46], as well as competitive inhibition between IgG1 and IgG2 subclasses [47]. These types of potential systemic errors were avoided in this work by using constant concentrations of purified serum isotypes for analysis.

In this work, COVID-19 disease severity was correlated with antibody isotype binding to a panel of antigens, and the selected antigen panels did not always coincide between antibody isotypes. Antibodies against multiple antigens presented by the pathogen are developed in the humoral immune response. Antibody ‘level’ or overall concentration in the blood, typically of IgG, has been associated with disease severity, in addition to level or intensity of IgG and IgA binding to various NP, S protein segments or RBD antigens [48]. Siracusano, *et al*. identified anti-S1 IgA binding early after PFSO as a strong clinical marker to discriminate the clinical course of COVID-19 disease, with higher binding associated with more severe illness [24]. Zervou, *et al*. also correlated high levels of serum IgA binding intensity against NP with severe disease [23]. Thus, it is feasible that using antibody isotype binding to panels of antigens can help to predict disease severity for clinical care and intervention.

Serological assays detecting specific serum antibody binding to various viral antigens have been developed as diagnostics for SARS-CoV-2 infections. Nonetheless, assays relying on a single antigen target can sacrifice sensitivity to improve specificity and avoid false positives or negatives. Regardless of thresholds imposed to avoid false positives, the lower antibody responses to certain antigens occurring earlier in infection may result in false negatives, and false positives can result from cross-reactivity due to previous exposure to sequentially similar antigens. For example, conserved SARS-CoV-1 and SARS-CoV-2 NP N-terminal domain residues are highly similar to endemic coronaviruses which cause the common cold (OC43, HKU1, NL63, and 229E) [22, 49]. The moderate-high binding intensity of pre-pandemic serum IgG, IgA and IgM to NP Ecoli and low-moderate binding to NP HEK and NP Sf21 (Figs. 2-5 and Table S3) observed in this work and others is due to cross-reactivity with the NP of endemic coronaviruses [22, 49, 50]. This cross-reactivity argues against the use of full length NP as a target antigen in any serological or diagnostic assays due to the high risk of false positives, either from previous exposure to, or current infection with, endemic coronaviruses. Indeed this has prompted the development and use of different SARS-CoV-2 NP fragments for greater specificity [22, 50]. Thus, reliance on a single antigen serological assay for diagnostic or monitoring is not advisable, particularly when antibodies against various antigens can develop at different times during the course of infection. For example, anti-S IgG typically appears after anti-NP IgG during infection, perhaps due to the greater abundance of NP [51] or perhaps the early anti-NP antibody response is anamnestic from previous exposure to structurally similar endemic coronaviruses NPs [22]. In addition to varying intensities of immune response across individuals with different disease severity demonstrated in this work and others [23, 51], it has also been shown that not all individuals develop antibodies against the same viral antigens [21]. Hence multiplexed assay formats presenting a panel of viral antigens are more desirable to accurately ascertain individual immune responses.

In this work, NP and S antigen glycosylation had a significant impact on antibody recognition and binding, with S glycosylation generally increasing and NP glycosylation decreasing antibody binding. The extensive S protein glycosylation plays critical roles in strong S protein binding to the host ACE2 receptor, including via glycan-glycan interactions, and contributes to immune evasion by shielding the protein backbone [9, 52]. Amanat, *et al*. reported higher IgG binding to RBD and S proteins expressed in the human-derived Expi293F compared to those expressed in insect cells [53] while Jiang, *et al*., suggested that the use of mammalian cell-expressed S1 protein in a multiplex assay format had higher specificity compared to *E. coli*-expressed S1 [28]. *E. coli*-expressed non-glycosylated RBD protein was previously demonstrated to elicit approximately 7-fold lower binding by immunised rat serum IgG at 5 μg/mL in an ELISA compared to glycosylated RBD expressed in insect (Sf21) and HEK cells [40]. At lower concentrations, binding to *E. coli*-expressed RBD was below the assay threshold. In addition, much weaker binding affinity of *E. coli*-expressed RBD to the ACE2 receptor was observed in comparison to insect- and HEK-expressed RBD binding (1.21 × 10^−6^ M versus 7.49 × 10^−9^ M and 5.39 × 10^−10^ M, respectively) [40]. These observations emphasise the importance of the use of appropriately glycosylated antigens for vaccine design and in serological assays for increased accuracy. With this in mind, the use of HEK-produced antigens is probably the most relevant to *in vivo* infection as HEK are human-derived cells and the produced antigen glycosylation will be the most human-like of the current commonly used commercially available antigen production systems of HEK and insect cells. However, in this work the S1 antigen produced in Sf21 insect cells (S1 Sf21) demonstrated significant association between COVID-19 disease severity and binding intensity for serum IgG, IgA and IgM (and S1 Full HEK for IgA and IgM). For selecting optimal expression systems, it must be noted that *in vivo* glycosylation of the produced viral proteins will vary depending on the individual and many factors including their blood type, biological sex, health status, and the relative stress on the infected cell producing the viral proteins.

Variations in the relative binding to the SARS-CoV-2 antigens were apparent for individuals with increasing time PFSO, and binding of specific isotypes to antigens even varied substantially in short time periods (e.g. 2 days). Not enough time points were collected in this work to ascertain when maximum antibody isotype binding to the antigens were reached for each patient, but there was a trend of earlier maximum binding for patients with milder disease (earlier than 30 days PFSO) and later maximum binding for patients with severe disease (30 days PFSO or later). This trend would have to be tested for greater confidence with larger patient numbers and more frequent sample points. Several of the same patient sera used in this study were also previously assessed in ELISA format by De Marco Verissimo *et al*. for changes in IgG binding to Npro Ecoli and S2 Frag Ecoli over time PFSO [21]. In agreement with De Marco Verissimo *et al*., binding to the same samples in this work increased over time except for three patient samples, which decreased over time in this work instead of increased as previously reported (compare patients 203-0015, 203-0023 and 203-0077 in Fig. S8 with Fig. 7 in [21]). However, later time points were used in this work for the same patients compared to earlier time points in De Marco Verissimo *et al*.’s study, which correlates with the decline in IgG binding over longer periods.

Total serum antibody reaches its maximum concentration 3 to 5 weeks PFSO [2, 7], and Iyer *et al*. reported maximum concentration of anti-RBD IgG, IgA, and IgM in serum between 14 – 28 days PFSO using standard dilutions of serum and plasma in ELISAs [54]. It is important to note from data in this work that the maximum antibody isotype binding for particular antigens did not generally agree with the maximum serum concentration of the antibody isotype (Figs. 6-8, S6, S7, and Table S10). Total antibody concentration or titre in blood typically increases substantially post-infection from the normal healthy baseline level and the total antibody concentration slowly decreases with time after the pathogen has been cleared. Along with the decrease in the total antibody concentration in the blood, the relative amount or proportion of the pathogen-specific antibodies should decrease also, since the stimulating infecting pathogen antigens are removed. Use of a dilution of plasma or serum in other antigen binding assay formats (e.g. ELISA) reflects the expected decrease in the overall concentration of total serum IgA and IgM over time, both of which have been reported as below the limit of detection by 3 months PFSO [2, 16]. However, this method does not give information on the relative proportion of the serum antibody isotype of the total serum population which bind to specific antigens, or any changes in relative affinity to specific antigens, over time. Indeed, use of a dilution of plasma or serum as the source of antibody isotypes will likely bias longitudinal serological assays, with samples from longer periods PFSO likely to result in even lower sensitivity due to the overall decreased total antibody concentration in blood in addition to the expected lower proportion of antigen-specific antibodies from the total antibody pool. In this work, the absolute concentrations of antibody isotypes in serum decreased over time during convalescence as expected (Tables 2 and S10). Using purified antibody isotypes to avoid introducing potential artefacts, we also observed the expected decrease of the relative proportion of specific SARS-CoV-2 antigens IgGs binding in severe sera over longer-term convalescence (5 and 9 months) post-COVID-19 disease. However, the relative proportion of antigen-specific IgA binding remained the same over this longer period, while that of IgM decreased for S antigens but was either maintained or increased for NP antigens.

Our observation of maintained relative proportion of antigen-specific IgA and IgM over longer convalescence periods has not been noted previously to the best of our knowledge. Various antibody isotypes including IgG, IgA, and IgM can work synergistically against enveloped viruses [7, 55]. Co-ordinated anti-S IgM and IgG responses after vaccination were reported to have significantly better virus-neutralising activity and higher antibody levels [56] and IgG3 and IgM together were demonstrated to contribute up to 80% of neutralisation in convalescent plasma, despite making up only 12% of the total antibody mass [57]. In addition, Ruggiero *et al*. reported a pattern of persistent anti-S IgM in 21.6% of a Pfizer-BioNTech COVID-19 mRNA vaccinated cohort of healthcare workers who had suffered previous SARS-CoV-2 infection one year prior to vaccination (i.e. the anti-S IgM was present in serum pre-vaccination after one year) [56]. It is worth noting that the Ruggiero *et al*. study was performed in typical ELISA format, so the overall concentration of the anti-S IgM in serum must have been relatively high even after one year. Typically, IgM responses are thought to be short-lived and to disappear post-IgG response. Nevertheless, there have been previous reports of antigen-specific memory IgM B cells post-infection including viral influenza and intracellular bacterial pathogens that persist for a lifetime [58-60]. Interestingly in this work, the IgM binding intensity to the S proteins declined in the two longer-term convalescent patients at 5 and 9 months, while IgM binding to NP proteins remained the same or even increased slightly (Fig. 8E and F). This could be because of previous exposure to the sequentially similar endemic coronaviruses NP proteins leading to an anamnestic response but different S proteins stimulating a canonical immune response. Alternatively, this may indicate the stimulation of different immune responses by the S and NP antigens leading to different immunological memory persistence. Our observations are consistent with a previous study which reported that of the very few COVID-19 patient samples tested that were IgM-positive, most of these were anti-NP IgM [48]. As SARS-CoV-2 may induce immunologically valuable IgM plasmablasts, it will be important to accurately determine the extent of SARS-CoV-2 antigen-specific IgM persistence across the population of different disease severities using a sensitive platform and methodology such as analysis of purified total IgM to help in formulating longer-lasting protective strategies.

Secretory IgA (mainly dimeric) plays a protective role as a component on mucosal surfaces but its link to (mainly monomeric) serum IgA is as yet unknown. Serum IgA was shown to play the dominant role in early neutralising SARS-CoV-2 response and were more potent than IgG in virus neutralisation assays, suggesting that serum IgA may play a more important role than IgG in early infection [3, 24]. Nonetheless, IgG responses are currently the most monitored post-vaccination and convalescence, with less attention paid to IgA and IgM responses. Recently, serum IgA was shown to increase phagocytosis of cancer cells by neutrophils compared to IgG [61], but it is not known if IgA can do the same for SARS-CoV-2 infected cells. A limitation of this study was that the availability of longer-term follow-up serum samples was limited to two patients who had suffered from severe COVID-19 disease. Thus, the observation of persistence of the proportion of IgA in severe patients after a longer convalescence period is of interest for further investigation with larger patient numbers and sampling from patients with different disease severities.

A robust RBD-binding IgG serum concentration has been reported in response to the Pfizer-BioNTech COVID-19 mRNA vaccine, greater in patients at 21 days after the first dose and 8-50-fold greater after the second dose compared to that of COVID-19 convalescent patients at least 14 days after RT-qPCR-confirmed infection [62]. In agreement, intense V1 and V2 IgG binding was noted 30 days after the second vaccine dose (Fig. 9). In contrast, the magnitude of IgG antigen binding intensity did not exceed that of convalescent patients with severe disease up to 33 days PFSO (Figs. 6 and 8) nor moderate disease up to 12 days PFSO (Fig. 7A), and was only approximately 20-30% greater than mild disease up to 34 days PFSO (Fig. 7B). This discrepancy in response magnitude is probably related to differences in assay type and in the serum sample preparation. The antigen microarray used in this work is a more sensitive platform than traditional ELISA-type assays and the purified isotype serum antibodies were assessed at consistent antibody isotype concentrations rather than the more typical dilution of whole serum and relying on specific isotype antibody detection. Both considerations can introduce their own biases, typically decreasing sensitivity.

Anti-S and anti-RBD IgG responses post-mRNA vaccination have been reported to be similar to that of SARS-CoV-2 infection [63-65], in agreement with our data. However in this work vaccinated donors V1 and V2 clearly demonstrated a lack of IgM or IgA response to S proteins 30 days after their second vaccine dose, in contrast to the detectable and robust IgM and IgA responses observed in COVID-19 convalescent patients. In a study of longitudinal IgG and IgM response in a large cohort of healthcare workers post-Pfizer-BioNTech COVID-19 mRN A vaccination, 77.4% of naïve vaccinees demonstrated non-canonical immune responses, with 36.1% developing IgG but no IgM response and 41.3% developing IgM after IgG [56]. Those with anti-S protein IgM positive sera demonstrated higher pseudovirus neutralisation titres in comparison to individuals who were IgM negative. Meanwhile, serum IgA responses post-mRNA vaccination were reported as variable and low in naïve individuals [63-65]. After two doses of mRNA vaccine only 41% of 107 tested long-term care home workers tested positive for anti-S IgA and 20% for anti-RBD IgA, with IgA titres significantly lower than COVID-19 convalescent patients at a similar time-point [63]. A recent study of a larger cohort demonstrated that mRNA vaccinated individuals who had been previously infected with SARS-CoV-2 produced higher IgG, IgA, and IgM responses against S antigens compared to naïve individuals, and antibody neutralisation capacity was also higher in pre-exposed compared to naïve individuals [65].

The lack of or very low serum IgA and IgM response to S proteins in the vaccinated patients is potentially concerning from the perspective of neutralising an active infection and maintaining long-term protection in the population. IgM is typically associated with early virus response pre-Ig class switching and IgA with interfering with pathogen transmission due to its abundance at mucosal sites. Further, a durable IgA response post-vaccination is associated with protection from subsequent SARS-CoV-2 infection [63]. In addition to our data, these reports highlight the importance of additionally targeting IgM and IgA responses in SARS-CoV-2 vaccine formulations and schedules for longer lasting effective protection.

## 5. Conclusions

In conclusion, the development and use of a sensitive multiplexed SARS-CoV-2 antigen microarray using purified serum antibody isotypes to assess and quantify binding allowed us to observe the confounding effect of the use of diluted whole serum on the pattern and quantity of antibody binding. Use of the more accurate purified antibody isotypes permitted correlation of isotype binding with panels of various antigens with COVID-19 disease severity, which may be useful for patient therapy and clinical management strategies. We observed that antigen glycosylation influenced the binding of IgG, IgA, and IgM, with S glycosylation generally increasing and NP glycosylation decreasing antibody binding. Finally, the relative proportion of total serum IgA binding to all SARS-CoV-2 antigens remained the same over longer convalescence periods, up to 5 and 9 months, while the relative proportion of total serum IgG binding declined substantially as expected. The relative proportion of total serum IgM binding over time dependence on the identity of antigen – relative binding stayed the same or slightly increased for NP antigens but decreased for S antigens. These data indicate that a long-term response of IgA and IgM binding to specific antigens might have an important role in longer-term protection, and a larger patient cohort should be studied for potential association.

## Data Availability

All data produced in the present work are contained in the manuscript and supplementary material

## Acknowledgements

MK acknowledges Science Foundation Ireland (SFI) COVID-19 Rapid Response Phase 2 Fund (grant number 20/COV/8477) for funding this study. BMcN thanks the Health Research Board for funding the National Irish COVID-19 Biobank (NICB). SD thanks SFI COVID-19 Rapid Response Phase 1 Fund (grant number 20/COV/0048).

