## Supplementary Information for "Correlation of patient serum IgG, IgA and IgM antigen binding with COVID-19 disease severity using multiplexed SARS-CoV-2 antigen microarray and maintained relative IgA and IgM antigen binding over time"

### Table of contents

**Fig. S1.** Purified SARS-CoV-2-positive patient (severe disease) serum IgG, IgA and IgM and the IgG-, IgM- and IgA-depleted flow through printed on a Nexterion® slide H surface and incubated with fluorescently labelled anti-isotype antibodies in different subarrays.

**Fig. S2.** Bar chart representing the binding intensity of His-tagged recombinant SARS-CoV-2 protein antigens to printed serum antibody isotypes purified from one SARS-CoV-2-positive patient (severe disease) serum sample.

**Fig. S3.** Antigen microarray print optimisation.

**Fig. S4.** Bar charts representing the binding intensity of diluted serum (1/100 in PBS-T) and purified serum antibody isotypes from a SARS-CoV-2-positive patient detected by fluorescently-labelled anti-isotype antibodies.

**Fig. S5.** Glycosylation profiling of SARS-CoV-2 antigens produced in mammalian (HEK) and insect (Sf21) cells.

**Fig. S6.** Dynamic binding of patient serum antibody isotypes over short term for severe COVID-19 disease.

**Fig. S7.** Dynamic binding of patient serum antibody isotypes over medium term for severe and mild COVID-19 disease.

**Fig. S8.** Variation in IgG binding to (**A**) Npro Full Ecoli and (**B**) S2Frag Ecoli for a subset of patient samples.

**Table S1.** Recombinant SARS-CoV-2 proteins used to construct the antigen (Ag) microarray.

**Table S2.** Lectins used for recombinant SARS-CoV-2 proteins glycoprofiling.

**Table S3.** Supplementary Excel file containing all antibody isotype binding to SARS-CoV-2 antigens from antigen microarray experiments and patient clinical data.

**Table S4.** Significance of correlation analysis for first sampling point serum IgG samples binding to antigens with low intensity compared to non-COVID-19 samples.

**Table S5.** Significance of correlation analysis for first sampling point serum IgG samples binding to antigens with high intensity compared to non-COVID-19 samples.

**Table S6.** Significance of correlation analysis for first sampling point serum IgA samples binding to antigens with low intensity compared to non-COVID-19 samples.

**Table S7.** Significance of correlation analysis for first sampling point serum IgA samples binding to antigens with high intensity compared to non-COVID-19 samples.

**Table S8.** Significance of correlation analysis for first sampling point serum IgM samples binding to antigens with low intensity compared to non-COVID-19 samples.

**Table S8.** Significance of correlation analysis for first sampling point serum IgM samples binding to antigens with high intensity compared to non-COVID-19 samples.

**Table S10.** Serum sample information including follow up samples.

A

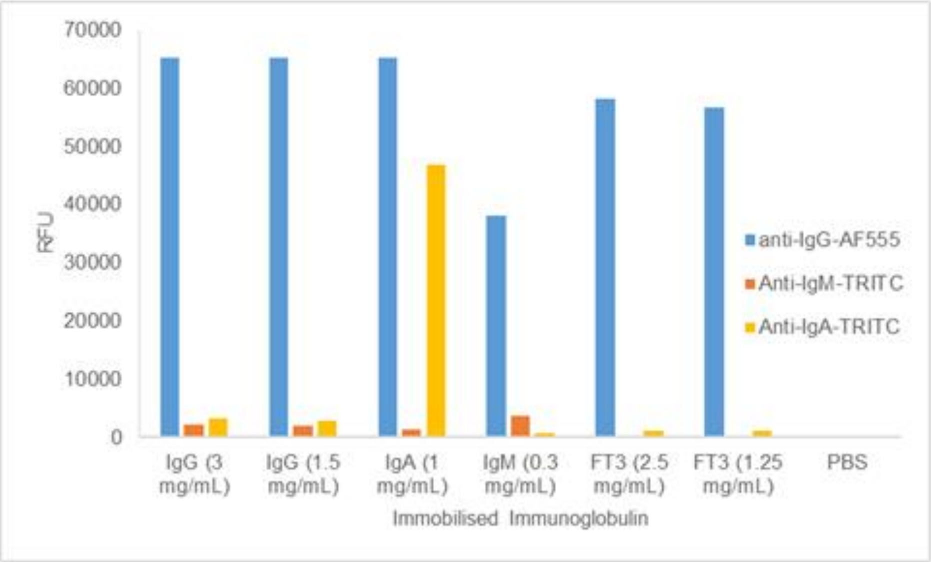


B

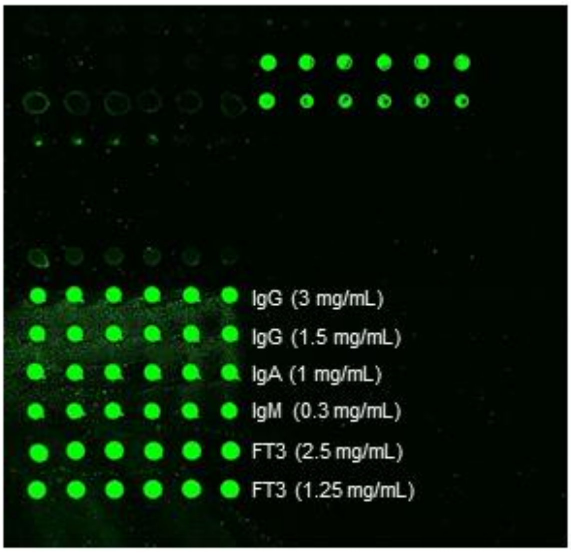


**Fig. S1.** Purified SARS-CoV-2-positive patient (severe disease) serum IgG, IgA and IgM and the IgG-, IgM- and IgA-depleted flow through printed on a Nexterion® slide H surface and incubated with fluorescently labelled anti-isotype antibodies in different subarrays. IgG was printed at 3 and 1.5 mg/mL, IgA at 1 mg/mL and IgM at 0.3 mg/mL and the flow through (FT3) at 2.5 and 1.25 mg/mL. (**A**) Bar chart representing the binding intensity of the fluorescently-labelled anti-isotype antibodies binding to immobilised IgG, IgA and IgM, incubated at 1, 2, and 1 µg/mL, respectively. (**B**) Scanned image of a subarray depicting the printed serum IgG, IgA, IgM and Ig-depleted FT3 incubated with fluorescently labelled anti-IgG antibody. The cross-reactivity of the anti-IgG antibody for IgA and IgM can be clearly observed and remaining IgGs were also detected in the Ig-depleted flow through FT3.

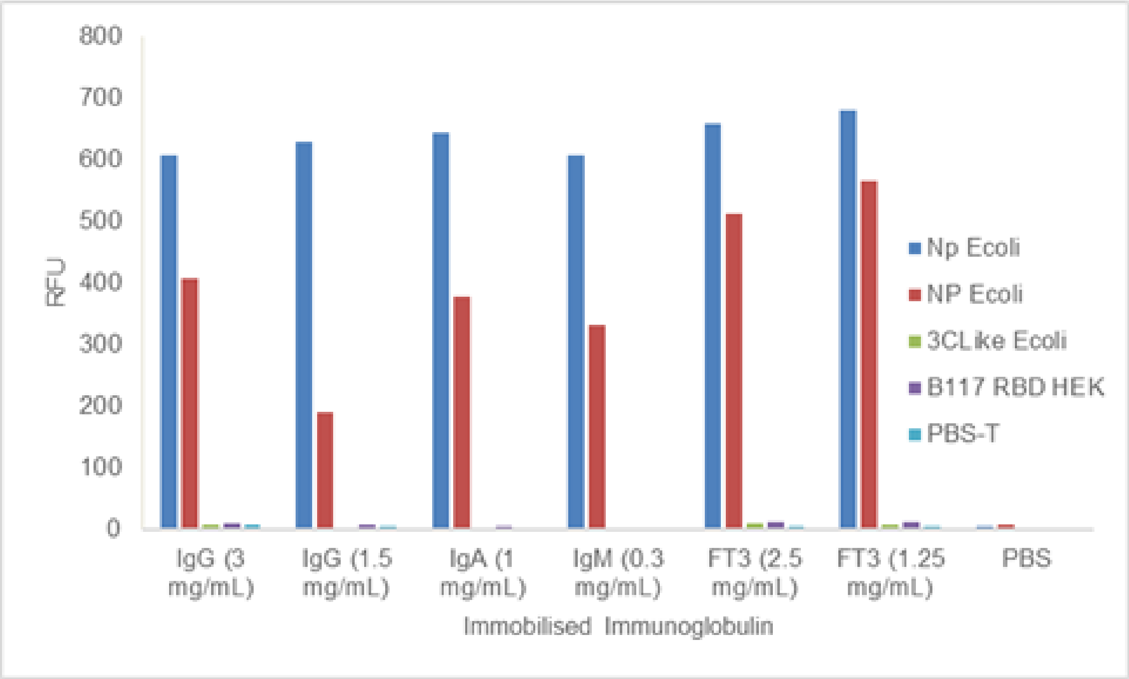


**Fig. S2.** Bar chart representing the binding intensity of His-tagged recombinant SARS-CoV-2 protein antigens to printed serum antibody isotypes purified from one SARS-CoV-2-positive patient (severe disease) serum sample. The serum antibody isotype microarray slides were incubated with a panel of antigens: NP Ecoli at 67 and 27 µg/mL, 3CLike Ecoli at 70 µg/mL and B117 RBD HEK at 17 µg/mL. Antigen binding was detected by incubation with fluorescently-labelled anti-His antibody at 1 µg/mL.

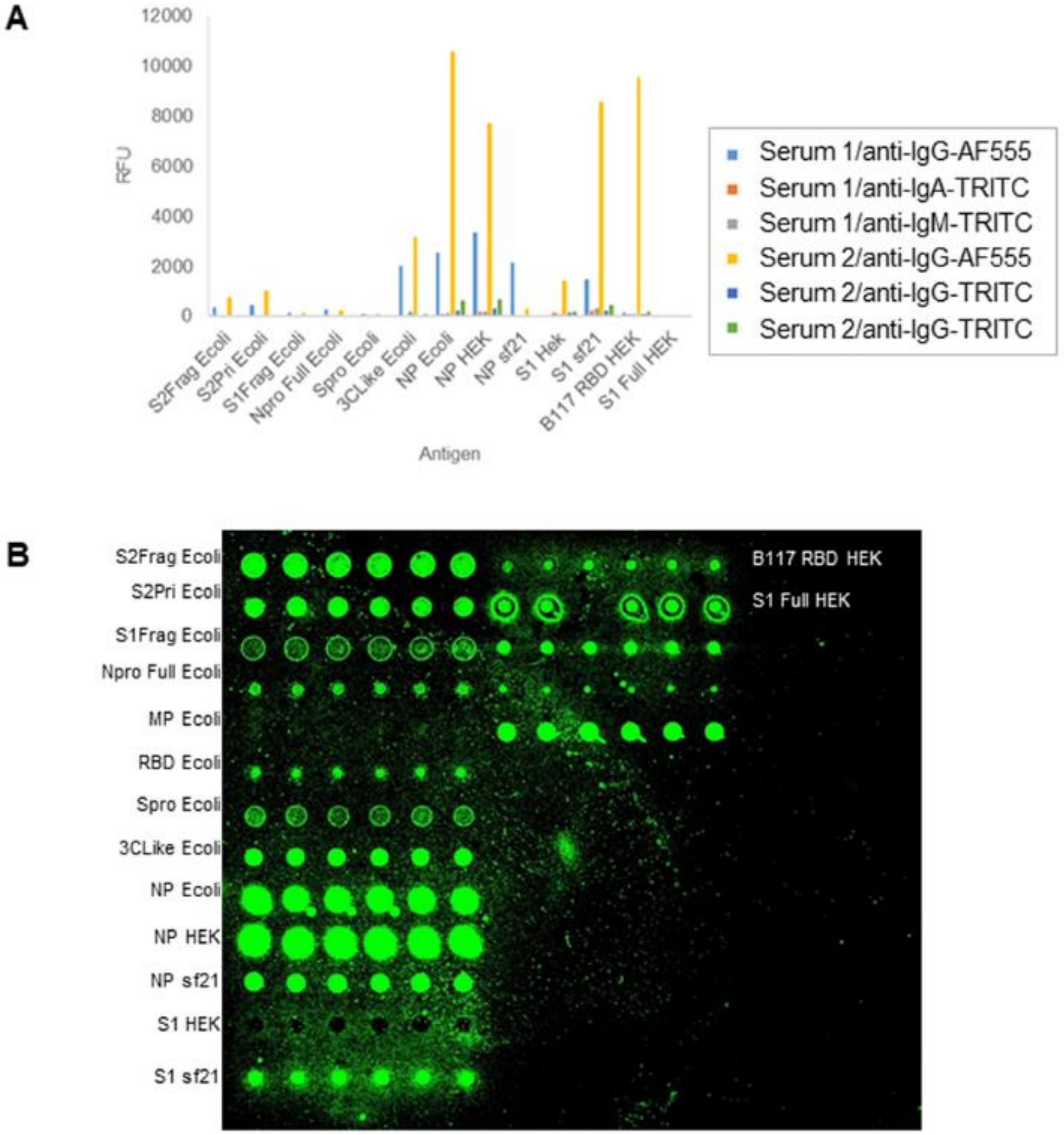


**Fig. S3.** Antigen microarray print optimisation. (**A**) Serum antibody isotype binding to SARS-CoV-2 antigens on microarray detected by either tetramethylrhodamine- (TRITC-) or AlexaFluor® 555- (AF555-)labelled anti-isotype antibodies. (**B**) Representative image of a subarray from an antigen microarray slide incubated with SARS-CoV-2-positive patient serum followed by detection with anti-IgG-AF555.

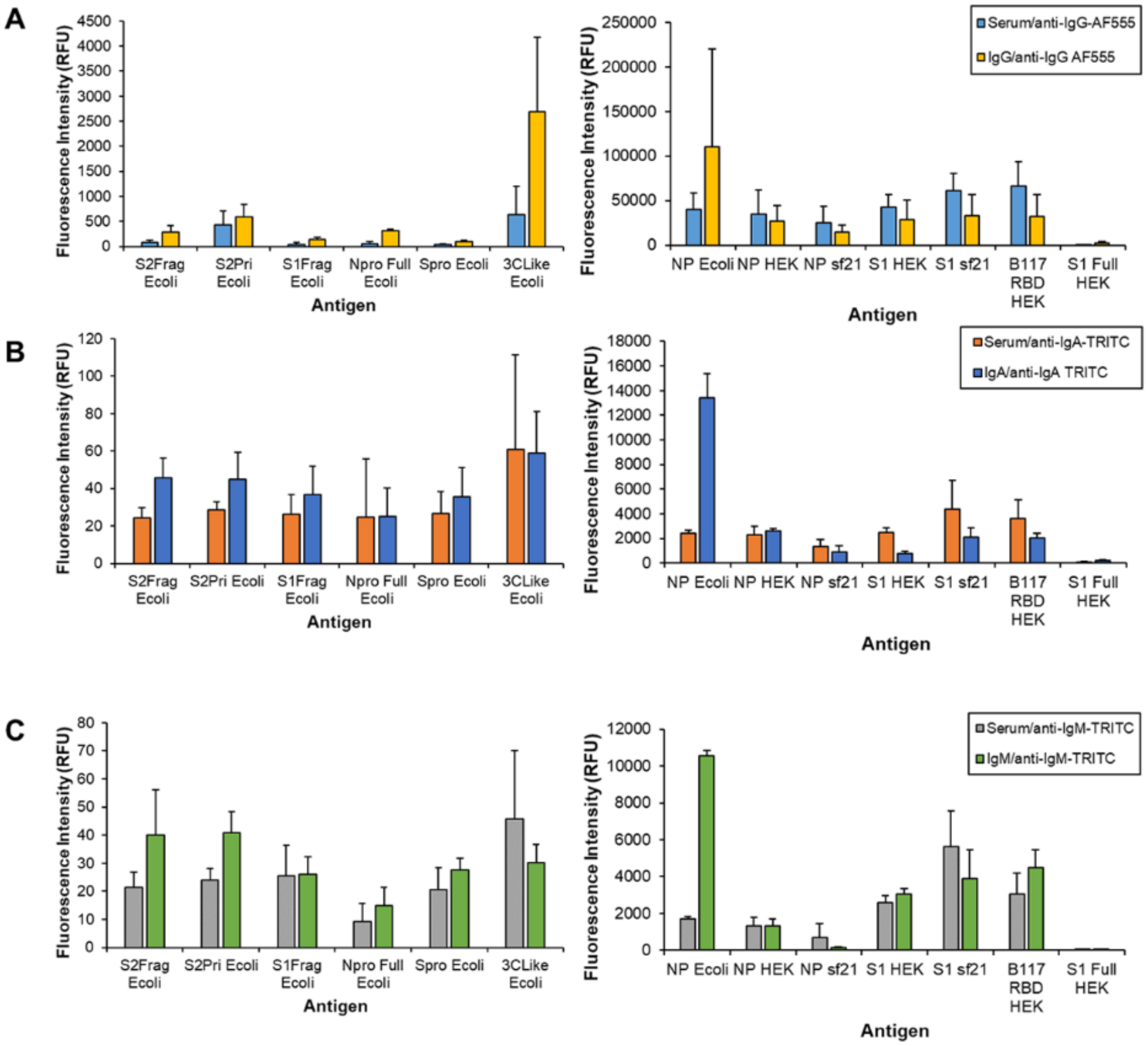


**Fig. S4.** Bar charts representing the binding intensity of diluted serum (1/100 in PBS-T) and purified serum antibody isotypes from a SARS-CoV-2-positive patient detected by fluorescently-labelled anti-isotype antibodies. Binding comparisons of (**A**) serum and purified serum IgG, (**B**) serum and purified IgA, and (**C**) serum and purified IgM. Comparisons of serum versus purified antibody isotype detection are shown across two charts to allow visualisation of lower serum antibody isotype binding to certain antigens.

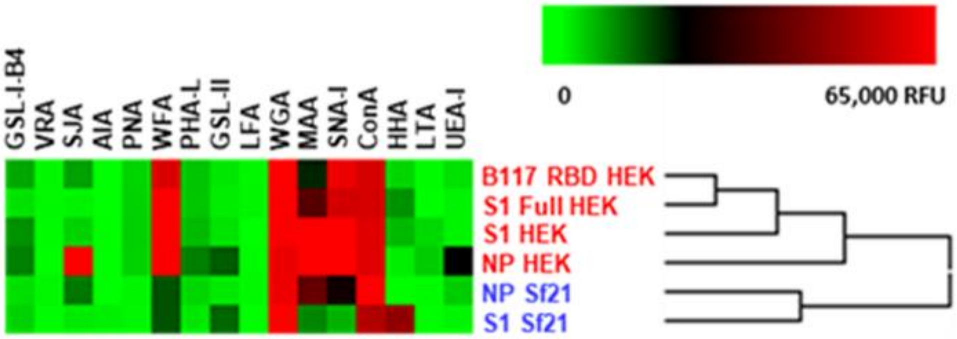


**Fig. S5.** Glycosylation profiling of SARS-CoV-2 antigens produced in mammalian (HEK) and insect (Sf21) cells. Unsupervised clustering of binding intensities of a panel of fluorescently-labelled lectins to recombinant SARS-CoV-2 proteins printed on the antigen microarray. The lectin LFA was non-functional in this format and was disregarded. Normalised data were subjected to unsupervised, Euclidean distance, complete linkage clustering.

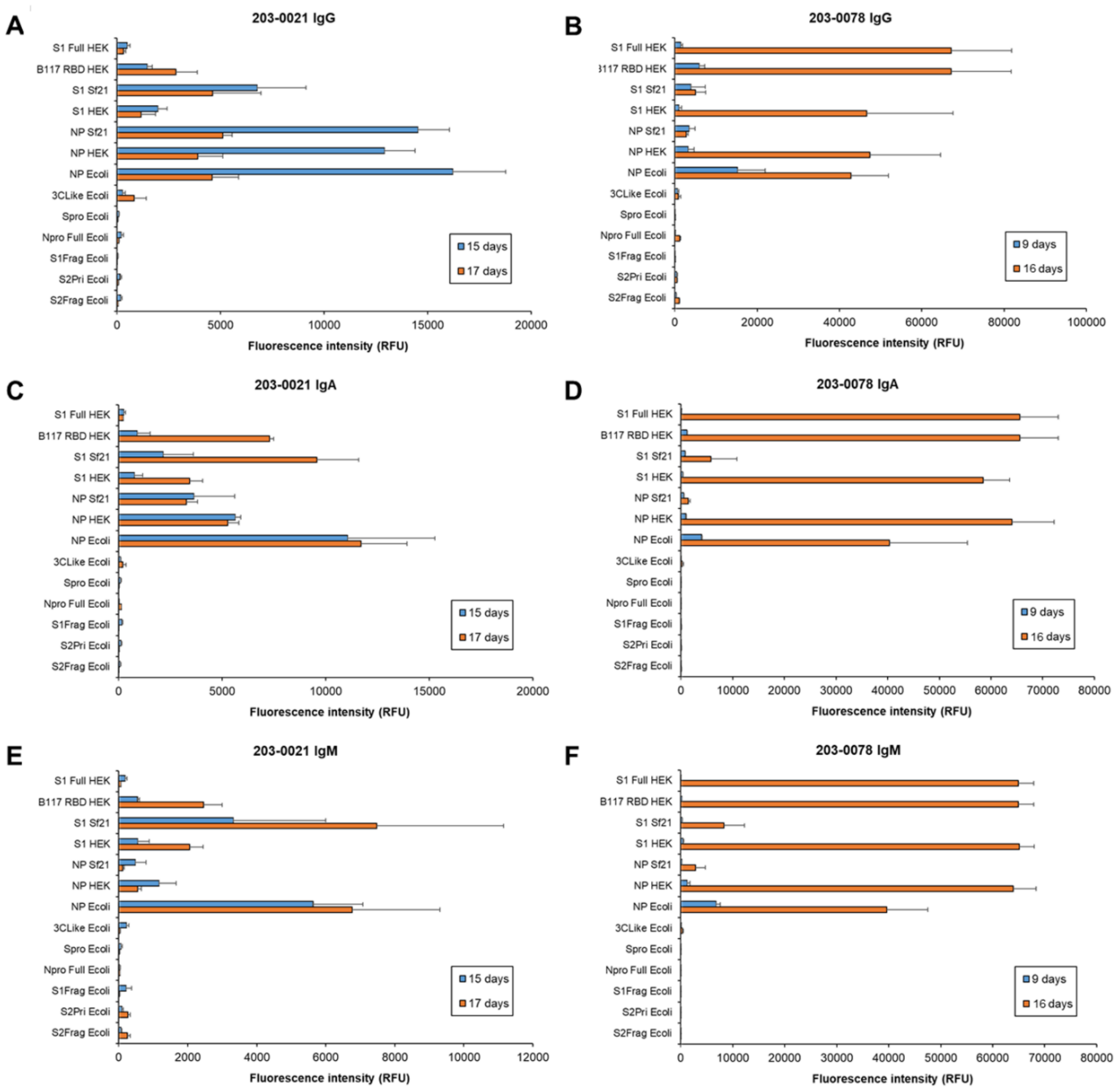


**Fig. S6.** Dynamic binding of patient serum antibody isotypes over short term for severe COVID-19 disease. Bar charts represent binding intensity data for serum (A,B) IgG, (C,D) IgA, and (E,F) IgM binding to SARS-CoV-2 antigens for patient 203-0021 (**A,C,D**; M, 61-65 years) and 203-0078 (**B,D,F**; F, 31-35 years) at 15 and 17 days, and 9 and 16 days post-first symptom onset, respectively.

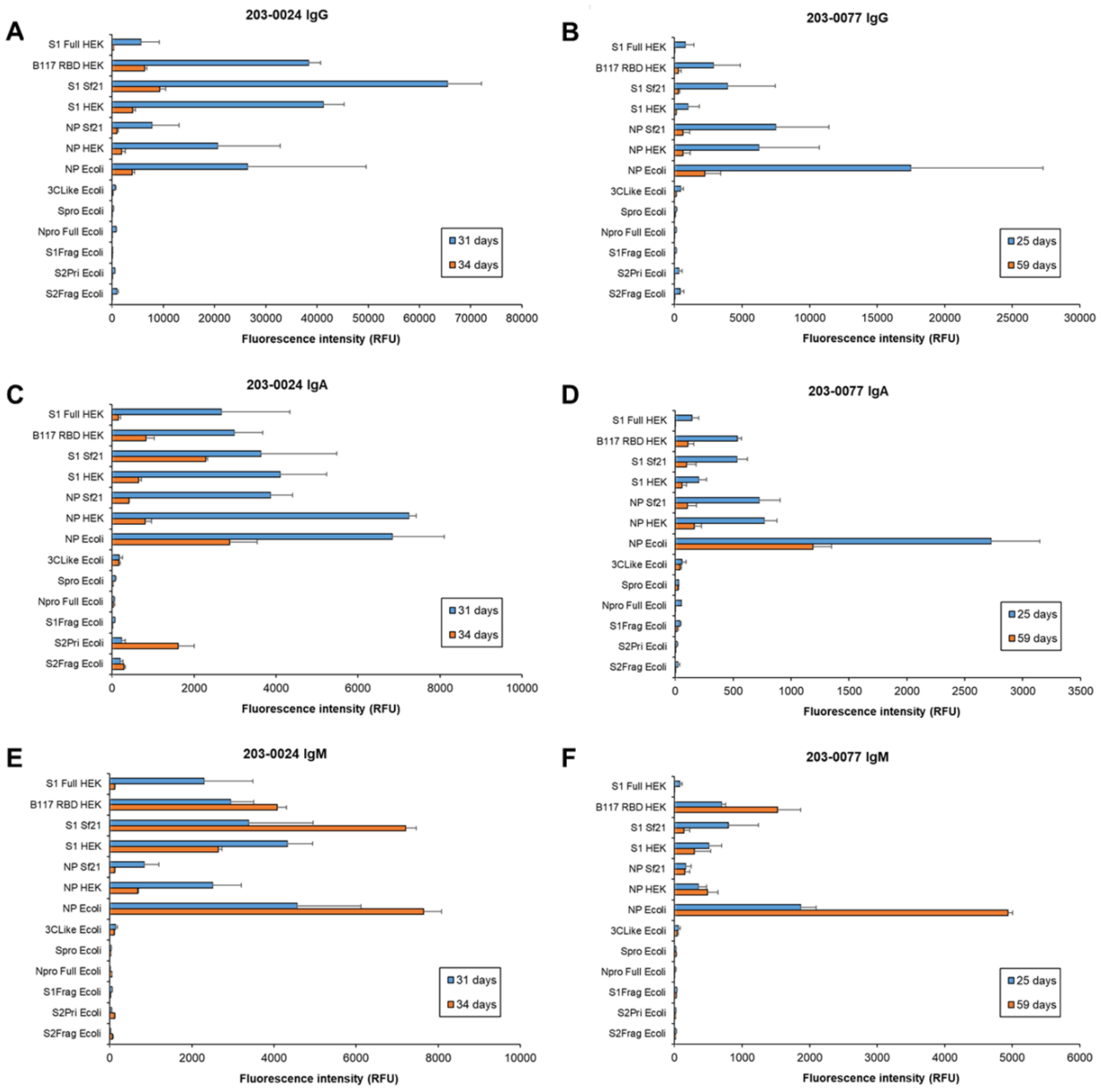


**Fig. S7.** Dynamic binding of patient serum antibody isotypes over medium term for severe and mild COVID-19 disease. Bar charts represent binding intensity data for serum (A,B) IgG, (C,D) IgA, and (E,F) IgM binding to SARS-CoV-2 antigens for patient 203-0024 (**A,C,D**; severe, F, 66-70 years) and 203-0078 (**B,D,F**; mild, M, 61-65 years) at 31 and 34 days, and 25 and 59 days post-first symptom onset, respectively.

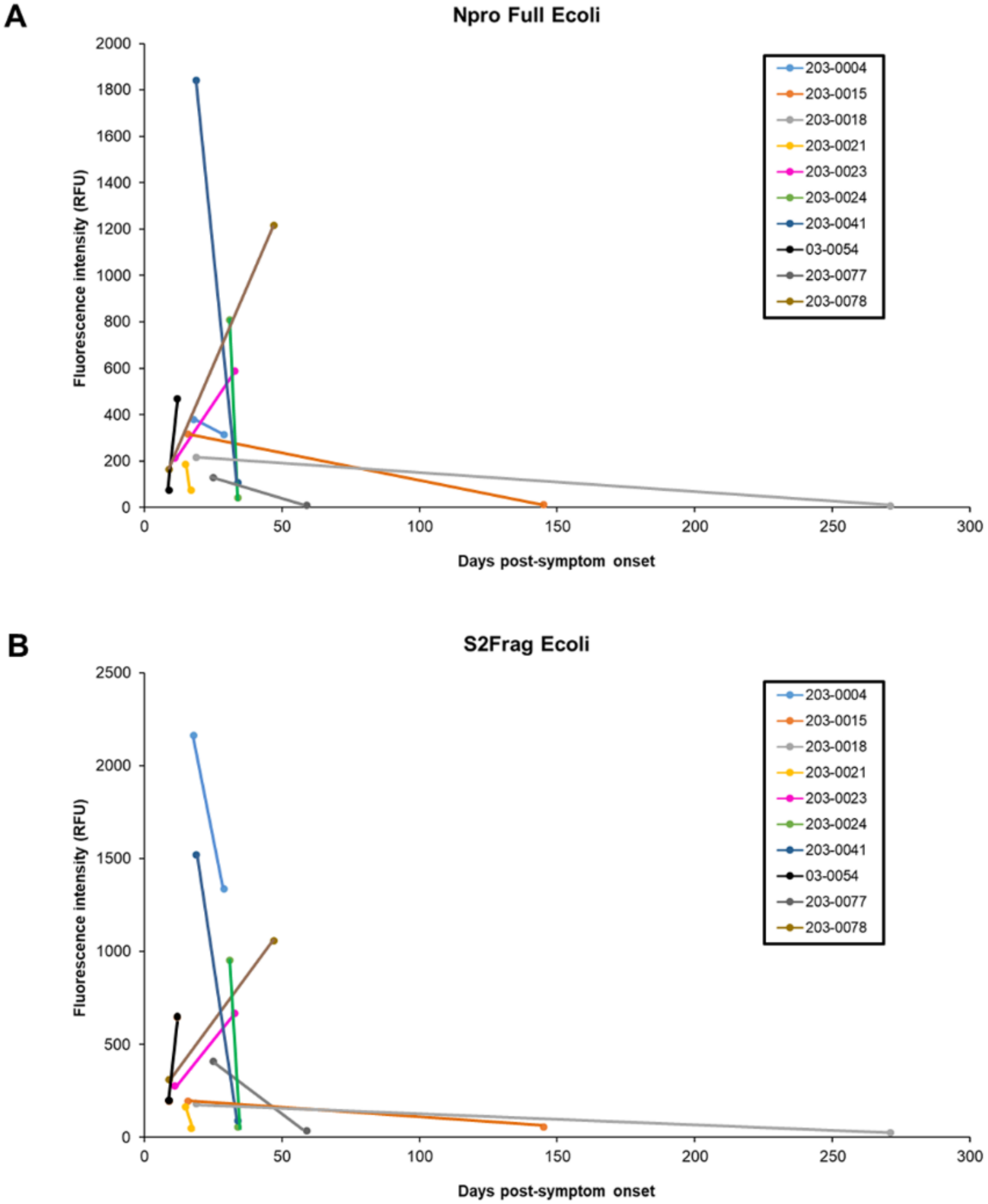


**Fig. S8.** Variation in IgG binding to (**A**) Npro Full Ecoli and (**B**) S2Frag Ecoli for a subset of patient samples.

**Table S1.** Recombinantly expressed SARS-CoV-2 proteins used to construct the antigen (Ag) microarray, the Ag code, approximate print concentration (mg/mL), molecular mass observed by SDS-PAGE, expression system, protein sequence expressed, buffer that the protein was supplied in, supplier, and catalogue number. MPL, Molecular Parasitology Lab; RB, RayBiotech; R&D, R&D Systems.

| Ag code | Protein Ag | Print conc | Mr (kDa) | Expression | Sequence | Buffer | Supplier | Cat. no. |
|---|---|---|---|---|---|---|---|---|
| Npro Full Ecoli | Nucleocapsid protein, full length | 0.06 | 50 | *E. coli* | 2-1269 | PBS + 500 mM imidazole | MPL | - |
| 3CLike Ecoli | 3C-like protease | 0.24 | 34 | *E. coli* | 1-306 | PBS | MPL | - |
| Spro Ecoli | Spike protein, full length | 0.12 | 135 | *E. coli* | 10-1282 | Potassium phosphate + 0.1% Sarkosyl, pH 7.4 | MPL | - |
| S1Frag Ecoli | Spike protein fragment 1 | 0.15 | 75 | *E. coli* | 10-686 | Potassium phosphate + 0.1% Sarkosyl, pH 7.4 | MPL | - |
| S2Frag Ecoli | Spike protein fragment 2 | 0.06 | 54 | *E. coli* | 687-1283 | Potassium phosphate + 0.1% Sarkosyl, pH 7.4 | MPL | - |
| S2Pri Ecoli | Spike protein fragment 2 prime region | 0.01 | 38 | *E. coli* | 816-1283 | PBS+0.05% Sarkosyl | MPL | - |
| NP Ecoli | Nucleocapsid protein | 0.95 | 50 | *E. coli* | Met1-Ala419 | PBS | RB | 230-01104 |
| NP HEK | Nucleocapsid protein | 0.6 | 50-60 | HEK293 | Met1-Ala419 | PBS | RB | 230-30164 |
| NP Sf21 | Nucleocapsid protein | 0.5 | 44-53 | Sf21 | Met1-Ala419 | PBS | R&D | 10474-CV |
| S1 HEK | S1 subunit protein | 0.5 | 106-121 | HEK293 | Val16-Pro681 | PBS with trehalose | R&D | 10569-CV |
| S1 Sf21 | S1 subunit protein | 0.2 | 78-92 | Sf21 | Val16-Pro681 | PBS with trehalose | R&D | 10522-CV |
| B117 RBD HEK | B.1.1.7 receptor binding domain | 0.25 | 34-38 | HEK293 | Arg319-Phe541 | PBS with trehalose | R&D | 10730-CV |
| S1 Full HEK | S1 subunit protein, full length | 0.45 | 120 | HEK293 | Val16-Gln690 | PBS | RB | 230-30161 |

| | | | | | | | | |
|---|---|---|---|---|---|---|---|---|
| MP Ecoli | Membrane glycoprotein, C-terminal | 0.17 | 15 | *E. coli* | Arg101-Gln222 | PBS + 200 mM imidazole | RB | 230-01124 |

**Table S2.** Lectins used for recombinant SARS-CoV-2 proteins glycoprofiling, their binding specificities and concentrations used (μg/mL).

| Lectin | Abbreviation | Major binding specificity | Conc. |
|---|---|---|---|
| *Griffonia simplicifolia* lectin | GSL-I-B4 | Terminal α-linked Gal | 10 |
| *Vigna radiate* lectin | VRA | Terminal α-linked Gal | 10 |
| *Artocarpus integrifolia* agglutinin | AIA | Gal-β-(1→3)-GalNAc (T-antigen), β-(1→6)-linked Gal, sialylation independent | 15 |
| *Arachis hypogaea* lectin | PNA | Gal-β-(1→3)-GalNAc (T-antigen) >GalNAc>Lac>Gal, terminal β-Gal) | 10 |
| *Sophora japonica* lectin | SJA | β-linked GalNAc>Gal | 10 |
| *Wisteria floribunda* agglutinin | WFA | GalNAc (GalNAc-α-(1→6)-Gal>GalNAc-α-(1→3)-R GalNAc (Forsmann antigen) >GalNAc>>Lac>Gal, chondroitin sulfate | 10 |
| *Phaseolus vulgaris* lectin | PHA-L | tri-, tetra-antennary β-Gal/Gal-β-(1→4)-GlcNAc | 10 |
| *Griffonia simplicifolia* lectin | GSL-II | GlcNAc | 10 |
| *Triticum vulgaris* lectin | WGA | NeuAc/GlcNAc | 10 |
| *Limax flavus* lectin | LFA | NeuAc/NeuGc | 10 |
| *Maackia amurensis* agglutinin | MAA | Neu-α-(2→3)-Gal = Gal-3-$SO_4$ >Lac | 15 |
| *Sambucus nigra* agglutinin I | SNA-I | Neu-α-(2→6)-Gal(NAc) >Lac, GalNAc >Gal | 10 |
| *Canavalia ensiformis* (jack bean lectin) | Con A | α-linked Man>Glc>GlcNAc | 10 |
| *Hippeastrum hybrid* lectin | HHA | Man-α-(1,3)-Man-α-(1,6)- | 10 |
| *Lotus tetragonolobus* lotus lectin | LTA | α-(1→3)-linked Fuc | 10 |
| *Ulex europaeus* agglutinin I | UEA-I | α-(1→2)-linked Fuc, H type 2 antigen | 10 |

**Table S4.** Significance of correlation analysis for first sampling point serum IgG samples binding to antigens with low intensity (3CLike Ecoli, Npro Full Ecoli, S1 Frag Ecoli, S2 Frag Ecoli, S2Pri Ecoli, and Spro Ecoli) compared to non-COVID-19 samples (NC). Bold *p* value indicates significant correlation.

| | | **IgG** | |
|---|---|---|---|
| *Predictors* | *Estimates* | *CI* | *p* |
| (Intercept) | 77.83 | 53.99 – 101.67 | **<0.001** |
| COVID.19 [Mild] | -36.13 | -73.61 – 1.35 | 0.058 |
| COVID.19 [Moderate] | 53.45 | 17.32 – 89.59 | **0.005** |
| COVID.19 [Severe] | -15.83 | -50.00 – 18.34 | 0.352 |
| Antigen [Npro Full Ecoli] | -39.05 | -64.36 – -13.74 | **0.003** |
| Antigen [S1Frag Ecoli] | -52.59 | -77.90 – -27.28 | **<0.001** |
| Antigen [S2Frag Ecoli] | -57.60 | -82.91 – -32.29 | **<0.001** |
| Antigen [S2Pri Ecoli] | -52.38 | -77.69 – -27.07 | **<0.001** |
| Antigen [Spro Ecoli] | -56.34 | -81.65 – -31.03 | **<0.001** |
| COVID.19 [Mild] * Antigen [Npro Full Ecoli] | 19.82 | -18.44 – 58.08 | 0.309 |
| COVID.19 [Moderate] *Antigen [Npro Full Ecoli] | -71.14 | -108.03 – -34.24 | **<0.001** |
| COVID.19 [Severe] *Antigen [Npro Full Ecoli] | 15.05 | -19.83 – 49.94 | 0.397 |
| COVID.19 [Mild] * Antigen [S1Frag Ecoli] | 80.40 | 42.14 – 118.67 | **<0.001** |
| COVID.19 [Moderate] *Antigen [S1Frag Ecoli] | -52.75 | -89.64 – -15.85 | **0.005** |
| COVID.19 [Severe] *Antigen [S1Frag Ecoli] | 44.99 | 10.10 – 79.88 | **0.012** |
| COVID.19 [Mild] * Antigen [S2Frag Ecoli] | 76.03 | 37.77 – 114.30 | **<0.001** |
| COVID.19 [Moderate] *Antigen [S2Frag Ecoli] | -45.17 | -82.07 – -8.28 | **0.017** |
| COVID.19 [Severe] *Antigen [S2Frag Ecoli] | 58.58 | 23.69 – 93.46 | **0.001** |
| COVID.19 [Mild] * Antigen [S2Pri Ecoli] | 63.15 | 24.88 – 101.41 | **0.001** |
| COVID.19 [Moderate] *Antigen [S2Pri Ecoli] | -56.01 | -92.91 – -19.12 | **0.003** |
| COVID.19 [Severe] *Antigen [S2Pri Ecoli] | 59.34 | 24.45 – 94.23 | **0.001** |
| COVID.19 [Mild] * Antigen [Spro Ecoli] | 63.97 | 25.71 – 102.24 | **0.001** |
| COVID.19 [Moderate] *Antigen [Spro Ecoli] | -44.13 | -81.02 – -7.23 | **0.019** |
| COVID.19 [Severe] *Antigen [Spro Ecoli] | 42.76 | 7.87 – 77.64 | **0.016** |
| $N_{id}$ | 34 | | |
| Observations | 612 | | |

**Table S5.** Significance of correlation analysis for first sampling point serum IgG samples binding to antigens with high intensity (B117 RBD HEK, NP Ecoli, NP HEK, NP Sf21, S1 Full HEK, S1 HEK, and S1 Sf21) compared to non-COVID-19 samples (NC). Bold *p* value indicates significant correlation.

| | | **IgG** | |
|---|---|---|---|
| *Predictors* | *Estimates* | *CI* | *p* |
| (Intercept) | 1229.48 | 431.16 – 2027.79 | **0.003** |
| COVID.19 [Mild] | -589.85 | -1845.15 – 665.45 | 0.345 |
| COVID.19 [Moderate] | 174.53 | -1035.83 – 1384.90 | 0.770 |
| COVID.19 [Severe] | 245.56 | -898.94 – 1390.06 | 0.664 |
| Antigen [NP Ecoli] | 4531.91 | 3914.78 – 5149.04 | **<0.001** |
| Antigen [NP HEK] | -201.15 | -818.28 – 415.99 | 0.522 |
| Antigen [NP Sf21] | -710.09 | -1327.22 – -92.96 | **0.024** |
| Antigen [S1 Full HEK] | -1041.49 | -1658.62 – -424.36 | **0.001** |
| Antigen [S1 Hek] | -994.09 | -1611.22 – -376.95 | **0.002** |
| Antigen [S1 Sf21] | -410.67 | -1027.80 – 206.46 | 0.192 |
| COVID.19 [Mild] *Antigen [NP Ecoli] | -2010.93 | -2943.94 – -1077.91 | **<0.001** |
| COVID.19 [Moderate] *Antigen [NP Ecoli] | -1984.75 | -2884.36 – -1085.13 | **<0.001** |
| COVID.19 [Severe] *Antigen [NP Ecoli] | -399.88 | -1250.54 – 450.78 | 0.356 |
| COVID.19 [Mild] * Antigen [NP HEK] | 432.73 | -500.29 – 1365.74 | 0.363 |
| COVID.19 [Moderate] *Antigen [NP HEK] | -137.27 | -1036.88 – 762.35 | 0.765 |
| COVID.19 [Severe] *Antigen [NP HEK] | 1381.22 | 530.56 – 2231.88 | **0.001** |
| COVID.19 [Mild] * Antigen [NP Sf21] | 516.01 | -417.01 – 1449.02 | 0.278 |
| COVID.19 [Moderate] *Antigen [NP Sf21] | 301.42 | -598.20 – 1201.04 | 0.511 |
| COVID.19 [Severe] *Antigen [NP Sf21] | 684.68 | -165.98 – 1535.34 | 0.114 |
| COVID.19 [Mild] * Antigen [S1 Full HEK] | 823.52 | -109.49 – 1756.54 | 0.084 |
| COVID.19 [Moderate] *Antigen [S1 Full HEK] | -304.15 | -1203.77 – 595.46 | 0.507 |
| COVID.19 [Severe] *Antigen [S1 Full HEK] | -55.03 | -905.69 – 795.62 | 0.899 |
| COVID.19 [Mild] * Antigen [S1 HEK] | 1081.81 | 148.80 – 2014.83 | **0.023** |
| COVID.19 [Moderate] *Antigen [S1 HEK] | 94.92 | -804.70 – 994.53 | 0.836 |
| COVID.19 [Severe] *Antigen [S1 HEK] | 571.77 | -278.89 – 1422.42 | 0.187 |
| COVID.19 [Mild] * Antigen [S1 Sf21] | 563.84 | -369.18 – 1496.85 | 0.236 |

| | | | |
|---|---|---|---|
| COVID.19 [Moderate] *Antigen [S1 Sf21] | 1187.56 | 287.95 – 2087.18 | **0.010** |
| COVID.19 [Severe] *Antigen [S1 Sf21] | 1536.96 | 686.30 – 2387.62 | **<0.001** |
| N $_{id}$ | 34 | | |
| Observations | 714 | | |

**Table S6.** Significance of correlation analysis for first sampling point serum IgA samples binding to antigens with low intensity (3CLike Ecoli, Npro Full Ecoli, S1 Frag Ecoli, S2 Frag Ecoli, S2Pri Ecoli, and Spro Ecoli) compared to non-COVID-19 (NC) samples. Bold *p* value indicates significant correlation.

| | | **IgA** | |
|---|---|---|---|
| *Predictors* | *Estimates* | *CI* | *p* |
| (Intercept) | 483.97 | 192.70 – 775.23 | **0.001** |
| COVID.19 [Mild] | 102.62 | -355.23 – 560.47 | 0.650 |
| COVID.19 [Moderate] | 995.78 | 554.32 – 1437.24 | **<0.001** |
| COVID.19 [Severe] | 0.29 | -417.15 – 417.72 | 0.999 |
| Antigen [Npro Full Ecoli] | -372.02 | -682.44 – -61.59 | **0.019** |
| Antigen [S1Frag Ecoli] | -431.12 | -741.55 – -120.70 | **0.007** |
| Antigen [S2Frag Ecoli] | -287.50 | -597.92 – 22.93 | 0.069 |
| Antigen [S2Pri Ecoli] | -283.09 | -593.52 – 27.33 | 0.074 |
| Antigen [Spro Ecoli] | -443.22 | -753.65 – -132.80 | **0.005** |
| COVID.19 [Mild] * Antigen [Npro Full Ecoli] | 206.66 | -262.66 – 675.98 | 0.387 |
| COVID.19 [Moderate] *Antigen [Npro Full Ecoli] | -905.28 | -1357.80 – -452.76 | **<0.001** |
| COVID.19 [Severe] *Antigen [Npro Full Ecoli] | 170.46 | -257.43 – 598.36 | 0.434 |
| COVID.19 [Mild] * Antigen [S1Frag Ecoli] | -67.87 | -537.19 – 401.45 | 0.776 |
| COVID.19 [Moderate] *Antigen [S1Frag Ecoli] | -916.57 | -1369.09 – -464.05 | **<0.001** |
| COVID.19 [Severe] *Antigen [S1Frag Ecoli] | 16.14 | -411.75 – 444.03 | 0.941 |
| COVID.19 [Mild] * Antigen [S2Frag Ecoli] | 176.54 | -292.78 – 645.86 | 0.460 |
| COVID.19 [Moderate] *Antigen [S2Frag Ecoli] | -785.90 | -1238.42 – -333.38 | **0.001** |
| COVID.19 [Severe] *Antigen [S2Frag Ecoli] | 282.46 | -145.43 – 710.35 | 0.195 |
| COVID.19 [Mild] *Antigen [S2Pri Ecoli] | 8.25 | -461.07 – 477.57 | 0.972 |
| COVID.19 [Moderate] *Antigen [S2Pri Ecoli] | -850.66 | -1303.18 – -398.15 | **<0.001** |
| COVID.19 [Severe] *Antigen [S2Pri Ecoli] | 505.82 | 77.93 – 933.71 | **0.021** |
| COVID.19 [Mild] *Antigen [Spro Ecoli] | -18.40 | -487.72 – 450.92 | 0.939 |
| COVID.19 [Moderate] *Antigen [Spro Ecoli] | -866.72 | -1319.24 – -414.20 | **<0.001** |
| COVID.19 [Severe] *Antigen [Spro Ecoli] | 78.89 | -349.00 – 506.78 | 0.717 |
| $N_{id}$ | 34 | | |
| Observations | 612 | | |

**Table S7.** Significance of correlation analysis for first sampling point serum IgA samples binding to antigens with high intensity (B117 RBD HEK, NP Ecoli, NP HEK, NP Sf21, S1 Full HEK, S1 HEK, and S1 Sf21) compared to non-COVID-19 samples (NC). Bold *p* value indicates significant correlation.

| | | **IgA** | |
|---|---|---|---|
| *Predictors* | *Estimates* | *CI* | *p* |
| (Intercept) | 2923.08 | -3469.64 – 9315.81 | 0.370 |
| COVID.19 [Mild] | 1262.03 | -8790.16 – 11314.22 | 0.799 |
| COVID.19 [Moderate] | 8141.82 | -1550.54 – 17834.17 | 0.097 |
| COVID.19 [Severe] | 15722.54 | 6557.67 – 24887.42 | **0.001** |
| Antigen [NP Ecoli] | 9101.42 | 4381.07 – 13821.77 | **<0.001** |
| Antigen [NP HEK] | 804.24 | -3916.11 – 5524.59 | 0.738 |
| Antigen [NP Sf21] | -744.45 | -5464.80 – 3975.90 | 0.757 |
| Antigen [S1 Full HEK] | -2210.84 | -6931.19 – 2509.51 | 0.358 |
| Antigen [S1 Hek] | -2322.92 | -7043.28 – 2397.43 | 0.334 |
| Antigen [S1 Sf21] | -568.70 | -5289.05 – 4151.65 | 0.813 |
| COVID.19 [Mild] * Antigen [NP Ecoli] | 1777.22 | -5359.28 – 8913.71 | 0.625 |
| COVID.19 [Moderate] *Antigen [NP Ecoli] | 6631.45 | -249.58 – 13512.49 | 0.059 |
| COVID.19 [Severe] *Antigen [NP Ecoli] | 2805.48 | -3701.08 – 9312.03 | 0.397 |
| COVID.19 [Mild] * Antigen [NP HEK] | 2800.65 | -4335.85 – 9937.15 | 0.441 |
| COVID.19 [Moderate] *Antigen [NP HEK] | 2122.99 | -4758.05 – 9004.02 | 0.545 |
| COVID.19 [Severe] *Antigen [NP HEK] | -4032.48 | -10539.03 – 2474.08 | 0.224 |
| COVID.19 [Mild] *Antigen [NP Sf21] | 1428.80 | -5707.70 – 8565.30 | 0.694 |
| COVID.19 [Moderate] *Antigen [NP Sf21] | 2052.18 | -4828.86 – 8933.21 | 0.558 |
| COVID.19 [Severe] *Antigen [NP Sf21] | -7715.67 | -14222.23 – -1209.12 | **0.020** |
| COVID.19 [Mild] *Antigen [S1 Full HEK] | -645.71 | -7782.21 – 6490.79 | 0.859 |
| COVID.19 [Moderate] *Antigen [S1 Full HEK] | -8431.77 | -15312.81 – -1550.74 | **0.016** |
| COVID.19 [Severe] *Antigen [S1 Full HEK] | -15285.98 | -21792.54 – -8779.43 | **<0.001** |
| COVID.19 [Mild] * Antigen [S1 HEK] | 4448.66 | -2687.84 – 11585.16 | 0.221 |
| COVID.19 [Moderate] *Antigen [S1 HEK] | -3845.54 | -10726.58 – 3035.49 | 0.273 |
| COVID.19 [Severe] *Antigen [S1 HEK] | -1546.90 | -8053.46 – 4959.65 | 0.641 |
| COVID.19 [Mild] *Antigen [S1 Sf21] | 4188.98 | -2947.52 – 11325.48 | 0.250 |

| | | | |
|---|---|---|---|
| COVID.19 [Moderate] *Antigen [S1 Sf21] | 3432.85 | -3448.19 – 10313.88 | 0.328 |
| COVID.19 [Severe] *Antigen [S1 Sf21] | 10480.62 | 3974.07 – 16987.18 | **0.002** |
| $N_{id}$ | 34 | | |
| Observations | 714 | | |

**Table S8.** Significance of correlation analysis for first sampling point serum IgM samples binding to antigens with low intensity (3CLike Ecoli, Npro Full Ecoli, S1 Frag Ecoli, S2 Frag Ecoli, S2Pri Ecoli, and Spro Ecoli) compared to non-COVID-19 samples (NC). Bold *p* value indicates significant correlation.

| | **IgM** | | |
|---|---|---|---|
| *Predictors* | *Estimates* | *CI* | *p* |
| (Intercept) | 82.21 | 61.80 – 102.62 | **<0.001** |
| COVID.19 [Mild] | 46.65 | 14.57 – 78.73 | **0.006** |
| COVID.19 [Moderate] | -31.78 | -62.71 – -0.85 | **0.044** |
| COVID.19 [Severe] | 17.79 | -11.47 – 47.04 | 0.224 |
| Antigen [Npro Full Ecoli] | -53.19 | -73.04 – -33.34 | **<0.001** |
| Antigen [S1Frag Ecoli] | -60.68 | -80.53 – -40.83 | **<0.001** |
| Antigen [S2Frag Ecoli] | -47.51 | -67.36 – -27.66 | **<0.001** |
| Antigen [S2Pri Ecoli] | -37.66 | -57.51 – -17.81 | **<0.001** |
| Antigen [Spro Ecoli] | -61.98 | -81.83 – -42.13 | **<0.001** |
| COVID.19 [Mild] * Antigen [Npro Full Ecoli] | -67.02 | -97.03 – -37.01 | **<0.001** |
| COVID.19 [Moderate] *Antigen [Npro Full Ecoli] | 15.23 | -13.70 – 44.17 | 0.302 |
| COVID.19 [Severe] *Antigen [Npro Full Ecoli] | -1.35 | -28.71 – 26.01 | 0.923 |
| COVID.19 [Mild] *Antigen [S1Frag Ecoli] | -8.29 | -38.30 – 21.72 | 0.588 |
| COVID.19 [Moderate] *Antigen [S1Frag Ecoli] | 34.59 | 5.65 – 63.53 | **0.019** |
| COVID.19 [Severe] *Antigen [S1Frag Ecoli] | 11.96 | -15.40 – 39.32 | 0.391 |
| COVID.19 [Mild] * Antigen [S2Frag Ecoli] | -47.83 | -77.84 – -17.82 | **0.002** |
| COVID.19 [Moderate] *Antigen [S2Frag Ecoli] | 37.51 | 8.57 – 66.45 | **0.011** |
| COVID.19 [Severe] *Antigen [S2Frag Ecoli] | -19.80 | -47.16 – 7.56 | 0.156 |
| COVID.19 [Mild] * Antigen [S2Pri Ecoli] | -52.67 | -82.69 – -22.66 | **0.001** |
| COVID.19 [Moderate] *Antigen [S2Pri Ecoli] | 38.44 | 9.50 – 67.37 | **0.009** |
| COVID.19 [Severe] *Antigen [S2Pri Ecoli] | -17.13 | -44.50 – 10.23 | 0.219 |
| COVID.19 [Mild] * Antigen [Spro Ecoli] | -26.12 | -56.13 – 3.89 | 0.088 |
| COVID.19 [Moderate] *Antigen [Spro Ecoli] | 36.00 | 7.06 – 64.94 | **0.015** |
| COVID.19 [Severe] *Antigen [Spro Ecoli] | -4.09 | -31.46 – 23.27 | 0.769 |
| $N_{id}$ | 34 | | |
| Observations | 612 | | |

**Table S9.** Significance of correlation analysis for first sampling point serum IgM samples binding to antigens with high intensity (B117 RBD HEK, NP Ecoli, NP HEK, NP Sf21, S1 Full HEK, S1 HEK, and S1 Sf21) compared to non-COVID-19 samples (NC). Bold *p* value indicates significant correlation.

| | | **IgM** | |
|---|---|---|---|
| *Predictors* | *Estimates* | *CI* | *p* |
| (Intercept) | 305.50 | -617.69 – 1228.69 | 0.516 |
| COVID.19 [Mild] | 647.38 | -804.29 – 2099.04 | 0.370 |
| COVID.19 [Moderate] | 1475.50 | 75.80 – 2875.20 | **0.039** |
| COVID.19 [Severe] | 1980.25 | 656.72 – 3303.77 | **0.005** |
| Antigen [NP Ecoli] | 3772.64 | 3062.98 – 4482.31 | **<0.001** |
| Antigen [NP HEK] | 266.87 | -442.80 – 976.53 | 0.461 |
| Antigen [NP Sf21] | -180.83 | -890.49 – 528.84 | 0.617 |
| Antigen [S1 Full HEK] | -217.19 | -926.86 – 492.47 | 0.548 |
| Antigen [S1 Hek] | -179.72 | -889.39 – 529.95 | 0.619 |
| Antigen [S1 Sf21] | -67.29 | -776.95 – 642.38 | 0.852 |
| COVID.19 [Mild] * Antigen [NP Ecoli] | -638.41 | -1711.32 – 434.51 | 0.243 |
| COVID.19 [Moderate] *Antigen [NP Ecoli] | -1286.73 | -2321.24 – -252.23 | **0.015** |
| COVID.19 [Severe] *Antigen [NP Ecoli] | -629.14 | -1607.35 – 349.07 | 0.207 |
| COVID.19 [Mild] * Antigen [NP HEK] | -479.05 | -1551.97 – 593.86 | 0.381 |
| COVID.19 [Moderate] *Antigen [NP HEK] | -1474.74 | -2509.25 – -440.23 | **0.005** |
| COVID.19 [Severe] *Antigen [NP HEK] | -214.26 | -1192.47 – 763.94 | 0.667 |
| COVID.19 [Mild] * Antigen [NP Sf21] | -653.36 | -1726.28 – 419.55 | 0.232 |
| COVID.19 [Moderate] *Antigen [NP Sf21] | -1231.12 | -2265.63 – -196.62 | **0.020** |
| COVID.19 [Severe] *Antigen [NP Sf21] | -1271.87 | -2250.07 – -293.66 | **0.011** |
| COVID.19 [Mild] * Antigen [S1 Full HEK] | -292.00 | -1364.92 – 780.91 | 0.593 |
| COVID.19 [Moderate] *Antigen [S1 Full HEK] | -1532.50 | -2567.01 – -497.99 | **0.004** |
| COVID.19 [Severe] *Antigen [S1 Full HEK] | -1772.80 | -2751.01 – -794.59 | **<0.001** |
| COVID.19 [Mild] * Antigen [S1 Hek] | 203.68 | -869.23 – 1276.60 | 0.709 |
| COVID.19 [Moderate] *Antigen [S1 Hek] | -410.73 | -1445.24 – 623.78 | 0.436 |
| COVID.19 [Severe] *Antigen [S1 Hek] | -375.87 | -1354.08 – 602.33 | 0.451 |
| COVID.19 [Mild] * Antigen [S1 Sf21] | 299.37 | -773.55 – 1372.28 | 0.584 |

| | | | |
|---|---|---|---|
| COVID.19 [Moderate] *Antigen [S1 Sf21] | 16.73 | -1017.78 – 1051.23 | 0.975 |
| COVID.19 [Severe] *Antigen [S1 Sf21] | 1549.98 | 571.77 – 2528.19 | **0.002** |
| $N_{id}$ | 34 | | |
| Observations | 714 | | |

**Table S10.** Serum sample information including follow up samples. Biobank code, COVID-19 disease status of individual at sampling time, days post-symptoms onset at sampling time, biological sex (M, male; F, female), patient age range at time of sampling, and serum IgG, IgA, and IgM concentration. N.d., not detectable; n.a., not assayed.

| Code | Disease | Days | Sex | Age range | Serum Ig conc (mg/mL) | | |
|---|---|---|---|---|---|---|---|
| | | | | | IgG | IgA | IgM |
| 203-0041-1 | Mild | 19 | M | 76-80 | 32.16 | 47.21 | 1.86 |
| 203-0041-3 | Mild | 34 | M | 76-80 | 5.14 | 6.53 | 0.24 |
| 203-0077-1 | Mild | 25 | M | 61-65 | 3.26 | 6.07 | 0.09 |
| 203-0077-3 | Mild | 59 | M | 61-65 | 1.13 | 1.63 | 0.16 |
| 203-0054-1 | Moderate | 9 | M | 46-50 | 13.26 | 4.07 | 0.31 |
| 203-0054-2 | Moderate | 12 | M | 46-50 | 113.33 | 45.92 | 4.49 |
| 203-0004-1 | Severe | 18 | M | 41-45 | 51.30 | 25.68 | 2.78 |
| 203-0004-2 | Severe | 29 | M | 41-45 | 8.11 | 2.20 | 0.27 |
| 203-0015-2 | Severe | 16 | M | 66-70 | 75.25 | 31.57 | 4.83 |
| 203-0015-4 | Severe | 145 | M | 66-70 | 1.13 | 1.62 | 0.17 |
| 203-0018-1 | Severe | 19 | M | 66-70 | 30.53 | 20.31 | 0.71 |
| 203-0018-4 | Severe | 271 | M | 66-70 | 1.00 | 0.52 | n.d. |
| 203-0021-1 | Severe | 15 | M | 61-65 | 50.99 | 12.93 | 3.71 |
| 203-0021-2 | Severe | 17 | M | 61-65 | 56.13 | 2.89 | 0.35 |
| 203-0023-1 | Severe | 11 | F | 56-60 | 58.52 | 48.17 | 1.79 |
| 203-0023-3 | Severe | 33 | F | 56-60 | 28.49 | 7.73 | 0.46 |
| 203-0024-1 | Severe | 31 | F | 66-70 | 83.84 | 29.13 | 3.61 |
| 203-0024-2 | Severe | 34 | F | 66-70 | n.a. | 9.08 | 1.55 |
| 203-0078-2 | Severe | 9 | F | 31-35 | 23.08 | 8.30 | 1.80 |
| 203-0078-3 | Severe | 16 | F | 31-35 | 66.61 | 19.34 | 4.03 |
